# The Effect of COVID-19 Hospitalisation on the Occurrence of Stroke: A Systematic Review and Meta-Analysis

**DOI:** 10.64898/2025.12.18.25342554

**Authors:** Khadija Sadiq, Raha Pazoki

## Abstract

**Background:** The relationship between COVID-19 hospitalisation and stroke occurrence remains incompletely understood, with reported incidence rates varying significantly across studies. This systematic review and meta-analysis aimed to comprehensively evaluate the association between COVID-19 hospitalisation and stroke occurrence, while examining key subgroup variations and potential risk factors.

**Methods:** A systematic search was conducted across PubMed, Scopus, and Web of Science databases from December 2019 to 2023. Studies reporting stroke occurrence in hospitalised COVID-19 patients were included. The Newcastle-Ottawa Scale was used for quality assessment. Random-effects meta-analysis was performed to calculate pooled odds ratios (OR) and risk ratios (RR) with 95% confidence intervals (CI).

**Results:** Nineteen studies met the inclusion criteria. The primary meta-analysis of 17 studies, comprising 98,297 patients, revealed a pooled stroke occurrence of 1% (95% CI: 1%,2%). A comparative analysis of three studies showed no significant difference in stroke risk between hospitalised COVID-19 patients and non-COVID-19 hospitalised controls (OR: 1.13, 95% CI: 0.49,2.65). However, within the COVID-19 hospitalised population, stroke risk was strongly associated with disease severity, including the need for mechanical ventilation (OR: 3.59, 95% CI: 2.21,5.83) and ICU admission (OR: 6.33, 95% CI: 4.95, 8.09). Significant pre-existing comorbidities included hypertension (OR: 2.35, 95% CI: 1.12,4.92) and atrial fibrillation (OR: 1.98, 95% CI: 1.18,3.31). Ischemic stroke was the predominant subtype, accounting for 81% of cases (95% CI: 73%,90%). Quality assessment of the 19 studies identified 9 as high quality and 10 as moderate quality.

**Conclusions:** This meta-analysis indicates that while the overall occurrence of stroke in hospitalised COVID-19 patients is approximately 1%, the risk is not significantly elevated compared to non-COVID-19 hospitalised patients. Instead, stroke occurrence is powerfully driven by the severity of the illness and the presence of traditional vascular risk factors. These findings underscore the critical need for vigilant neurological monitoring and targeted stroke prevention strategies, particularly for COVID-19 patients who require intensive care or mechanical ventilation.

## Introduction

### Global Burden and Clinical Significance

The convergence of COVID-19 and cerebrovascular disease represents one of the most significant challenges in contemporary medicine, building upon an already substantial global stroke burden. Prior to the pandemic, stroke claimed position as the second-leading cause of death globally, with the Global Burden of Disease Study 2019 (1) documenting 12.2 million annual incident cases and 101 million prevalent cases worldwide. This pre-existing burden was characterised by stark geographical disparities, with age-standardised mortality rates 3.6 times higher in low-income compared to high-income countries, reflecting profound inequities in prevention, access to care, and outcomes.

The emergence of SARS-CoV-2 introduced unprecedented complexity to this landscape through multiple pathogenic mechanisms. Early investigations by Klok et al. (2) revealed a striking 31% incidence of thrombotic complications in COVID-19 ICU patients despite standard prophylaxis, suggesting novel pathophysiological pathways. This observation was further substantiated by Beyrouti et al.’s documentation (3) of elevated D-dimer levels (>7000 μg/L) and positive antiphospholipid antibodies in COVID-19 stroke patients, indicating unique prothrombotic mechanisms distinct from traditional stroke pathways.

### Evolution of Understanding and Current Knowledge

The temporal progression of understanding the effect of COVID-19 hospitalisation on stroke occurrence has revealed increasingly complex patterns requiring systematic investigation. Initial reports from Wuhan by Mao et al. (4) documented acute cerebrovascular events in 5.7% of severe COVID-19 cases versus 0.8% in non-severe cases, establishing the first link between disease severity and stroke risk. This understanding evolved through distinct phases, each providing crucial insights. Early pandemic data suggested dramatically elevated stroke risk, with Merkler et al. (5) reporting 7.6-fold increased odds compared to influenza patients (95% CI: 2.3,25.2), predominantly featuring large vessel occlusions in younger patients without traditional risk factors.

Subsequent larger-scale investigations provided more nuanced understanding. Nannoni et al.’s (6) meta-analysis of 108,571 patients revealed a more precise cerebrovascular disease rate of 1.4% (95% CI: 1.0,1.9%), while identifying distinct prothrombotic profiles and mechanistic insights. This refinement culminated in Mortensen et al.’s (7) nationwide Danish registry study, demonstrating that while only 0.6% of acute ischemic stroke patients had concurrent SARS-CoV-2 infection, these cases presented with significantly more severe strokes (adjusted relative risk 1.93, 95% CI: 1.22,3.04) and higher 30-day mortality (adjusted relative risk 2.29, 95% CI: 1.19,4.39).

This evolution from early high estimates to more precise risk quantification reflects improved understanding and potentially changing virus-host interactions. The progression has revealed complex interactions between traditional vascular risk factors and COVID-19-specific mechanisms, while highlighting the importance of healthcare system factors in stroke detection and care delivery. These insights underscore the need for sophisticated analysis of emerging patterns and mechanisms in COVID-19-related stroke.

### Healthcare System Impact and Clinical Patterns

The pandemic’s impact on stroke care has been profound and quantifiably significant. Global registry data from Nogueira et al. (8), analysing 457 stroke centres across 70 countries, documented an 11.5% decline in stroke admissions during the pandemic’s first wave, with primary stroke centers and those with higher COVID-19 volumes experiencing steeper declines. This disruption occurred alongside evidence of compromised care metrics, with Srivastava et al. (9) demonstrating significantly extended treatment times in COVID-19 stroke patients: longer door-to-CT times (median 55 vs. 35 minutes, p<0.001), door-to-needle times (59 vs. 46 minutes, p<0.001), and door-to-endovascular therapy times (114 vs. 90 minutes, p=0.002).

The impact extended beyond timing metrics to clinical outcomes. Srivastava et al. (9) from The Get With The Guidelines-Stroke registry analysis revealed COVID-19 stroke patients had decreased odds of favourable discharge outcomes (OR 0.65, 95% CI: 0.52-0.81) and increased in-hospital mortality (OR 4.34, 95% CI: 3.48-5.40). Notably, Katz et al. (10) identified COVID-19 as the strongest independent risk factor for in-hospital stroke (OR 20.9, 95% CI: 10.4-42.2), with a striking shift toward in-hospital stroke onset (47.7% vs. 5.0%, p<0.001) compared to pre-pandemic patterns.

These findings highlight a dual challenge: reduced access to stroke care alongside increased complexity and poorer outcomes in COVID-19-associated strokes. The recovery patterns observed in later pandemic months (9.5% volume increase, May-June vs. March-April 2020) suggest healthcare systems’ adaptive capacity but underscore the need for robust stroke care protocols during infectious disease outbreaks.

### Pathophysiological Mechanisms and Clinical Phenotypes

The mechanisms underlying COVID-19-associated stroke represent a complex interplay of direct viral effects, inflammatory responses, and coagulation dysregulation. Systematic analyses by Nannoni et al. (6) revealed distinct pathogenic patterns, with COVID-19 stroke patients showing significantly elevated D-dimer levels (mean 9.2 ± 14.8 mg/L) and a high frequency of large vessel occlusions (OR 2.73, 95% CI: 1.63,4.57) compared to non-COVID-19 stroke patients. This prothrombotic state was further evidenced by Klok et al.’s (2) pivotal ICU study documenting a 31% incidence of thrombotic complications despite standard prophylaxis.

The clinical phenotype demonstrates several distinctive features. Yaghi et al. (11) reported a striking predominance of cryptogenic strokes in COVID-19 patients (65.6%) compared to both contemporary (30.4%, p=0.003) and historical controls (25.0%, p<0.001). Beyrouti et al. (3) identified a characteristic pattern of elevated antiphospholipid antibodies and extreme D-dimer elevation (>7000 μg/L), particularly in severe cases. Lodigiani et al. (12) observed that 50% of thromboembolic events occurred within 24 hours of admission, suggesting both immediate and delayed stroke mechanisms.

Three primary mechanistic pathways have emerged through systematic investigation, direct endothelial dysfunction through ACE2-mediated viral invasion, systemic inflammatory response leading to hypercoagulability and cardiac complications promoting thromboembolism. The predominance of large vessel occlusions and multi-territory infarcts, even in younger patients without traditional risk factors, suggests these mechanisms may operate synergistically, creating a unique stroke phenotype requiring specific therapeutic approaches (Tan et al. (13)).

### Temporal Evolution and Healthcare System Impact

The pandemic’s impact on stroke care delivery has evolved significantly since its onset. Nogueira et al. (8) documented an 11.5% decline in stroke admissions across 457 stroke centers globally during the pandemic’s first wave (95% CI: -11.7 to -11.3, p<0.0001), with primary stroke centers experiencing steeper declines than comprehensive centers. This disruption occurred alongside evidence of increased stroke severity in COVID-19 patients, as demonstrated by Srivastava et al. (9) through significantly extended treatment metrics: longer door-to-CT times (median 55 vs. 35 minutes, p<0.001) and door-to-needle times (59 vs. 46 minutes, p<0.001).

The temporal pattern of stroke occurrence has shown distinct phases. Early pandemic studies by Merkler et al. (5) suggested dramatically elevated risk (OR 7.6, 95% CI: 2.3,25.2) compared to influenza patients. However, more recent evidence from Mortensen et al. (7) revealed that while only 0.6% of acute ischemic stroke patients had concurrent SARS-CoV-2 infection, these cases presented with significantly more severe strokes (adjusted relative risk 1.93, 95% CI: 1.22,3.04) and higher mortality (aRR 2.29, 95% CI: 1.19,4.39).

### Critical Areas of Uncertainty

The existing evidence base, while substantial, presents several critical limitations that this investigation aims to address. Previous systematic reviews, including those by Tan et al. (13) and Nannoni et al. (6), were necessarily limited by the predominance of early pandemic data and inability to account for temporal changes in viral variants and treatment protocols.

COVID-19’s true impact on stroke occurrence remains uncertain, with rates varying from 0.6% (Mortensen et al. (7)) to 5.7% in severe cases (Mao et al. (4)).

Questions persist about healthcare system influences on stroke detection and the relationship between disease severity markers and stroke risk. While Klok et al. (2) found 31% thrombotic complications in ICU patients, the connection between mechanical ventilation and stroke occurrence needs clarification, including the role of respiratory severity markers and inflammatory indicators in predicting stroke risk.

Risk factor analysis reveals uncertainties in gender-specific stroke patterns, age distributions challenging traditional paradigms, and the impact of pre-existing conditions. Stroke pattern characterisation remains unclear, particularly regarding stroke subtypes and the mechanisms behind the high proportion of cryptogenic strokes reported by Yaghi et al. (11). The relationship between stroke timing and COVID-19 progression also requires investigation.

Contextual factors present additional uncertainties, including geographical variations in outcomes, pandemic phases’ impact on stroke detection, healthcare resource influences, and stroke pattern evolution across viral variants.

This systematic review and meta-analysis aims to quantify the global pooled occurrence of stroke in patients hospitalised with COVID-19. We seek to identify key demographic and clinical factors that influence stroke risk in this population.

## Methods

### Methodological Framework and Analytical Approach

Our methodological framework addresses the complexities of COVID-19-related stroke research through innovative meta-analytical approaches designed to overcome previously identified limitations and heterogeneity issues.

We implement a refined Newcastle-Ottawa Scale that specifically addresses pandemic-era research challenges, including variable follow-up periods and inconsistent disease severity reporting. This enables detailed examination of effect modifiers and subgroup analyses, crucial given the complex COVID-19 severity-stroke risk relationship identified by Merkler et al. (5).

The statistical methodology incorporates advanced proportion meta-analysis frameworks to handle diverse study designs while maintaining robust subgroup analysis capabilities. This approach integrates data from various healthcare settings and pandemic phases, building on Nogueira et al.’s (8) global registry findings.

Our framework evaluates mechanical ventilation’s impact on stroke occurrence (OR: 3.81, 95% CI: 2.68,5.42) while assessing demographic patterns, comorbidities, and healthcare system factors. This enables analysis of geographical variations and pandemic phase effects, providing insights into COVID-19 hospitalisation and stroke occurrence relationships.

### Search Strategy and Criteria

A comprehensive literature search was conducted using three major electronic databases including PubMed, Scopus, and Web of Science.

All searches were conducted using title and abstract filters, with appropriate adjustments made for the specific syntax requirements of each database (e.g., "Title/Abstract" for PubMed, "TS=" for Web of Science, and "TITLE-ABS-KEY" for Scopus). No language or date restrictions were applied to the searches to ensure maximum coverage of relevant literature.

To ensure comprehensive coverage of the available literature, additional search methods were employed. The reference lists of included studies and relevant review articles were manually screened using Endnote to identify additional eligible studies that may have been missed during the primary database searches. This process, known as backward citation tracking, helped to find studies that met the inclusion criteria but were not captured by the electronic search strategy.

However, hand-searching of conference proceedings and searching of clinical trial registries, such as ClinicalTrials.gov, were not conducted for this review. This limitation was acknowledged and considered when assessing the comprehensiveness of the search strategy and the potential for publication bias.

### Inclusion and Exclusion Criteria

This systematic review included primary research articles encompassing randomised controlled trials, cohort studies (both prospective and retrospective), case-control studies, cross-sectional studies, and observational studies published in English from December 2019 onwards. The primary population of interest comprised patients hospitalised with laboratory-confirmed COVID-19 infection, regardless of age or geographical location. Studies were required to include either patients who developed new stroke during hospitalisation or those with pre-existing stroke who experienced worsening symptom, new stroke-related complications, or progression of existing stroke conditions during their COVID-19 hospitalisation, with clear documentation of baseline stroke status on admission.

For inclusion, studies needed to report at least one of our primary outcomes, stroke incidence or prevalence in hospitalised COVID-19 patients, associated risk measures (odds ratio, relative risk, or hazard ratio), or timing of stroke onset relative to hospitalisation. Secondary outcomes of interest included stroke severity in hospital setting, length of hospital stay, ICU admission rates, in-hospital mortality, and TIA when reported alongside stroke outcomes. Methodologically, studies were required to provide clear documentation of both COVID-19 diagnosis (through PCR testing, antibody testing, or clear diagnostic criteria during hospitalisation) and stroke diagnosis (confirmed through hospital-based neuroimaging, in-hospital clinical assessments, or hospital-based biomarker analysis). Additionally, studies needed to document hospitalisation details including admission dates, length of stay, and level of care (general ward/ICU).

Studies were excluded if they were case reports, case series, systematic reviews, meta-analyses, opinion pieces, editorials, letters without original data, or conference abstracts. We also excluded studies that lacked clear hospitalisation data, mixed inpatient and outpatient data without clear separation, had unclear diagnostic criteria for either COVID-19 or stroke, or provided insufficient statistical analysis, or where the temporal relationship between hospitalisation and stroke was unclear. Studies focusing solely on outpatient outcomes, post-discharge outcomes, or COVID-19 vaccination were excluded, as were those reporting stroke occurrence before hospitalisation or primarily attributing stroke to pre-existing conditions. Studies not specifying hospital setting, animal studies, in vitro studies, and duplicate publications were also excluded. This comprehensive set of inclusion and exclusion criteria was established a priori to ensure systematic and unbiased study selection while maintaining methodological rigor, following PICO guidelines.

The screening and selection process was conducted by a single reviewer. The initial database searches identified 3,743 records (PubMed: 1,114; Scopus: 1,444; Web of Science: 1,185). After removing 1,481 duplicate records, 2,262 unique articles remained for screening. Of these, 2,073 records were excluded during initial screening based on title and abstract review. The remaining 189 full-text articles were assessed for retrieval, of which 2 were not retrievable. Among the 187 full-text articles assessed for eligibility, 168 were excluded for the following reasons: wrong study design (n=4), wrong outcomes with no stroke occurrence data (n=120), wrong patient population focusing on outpatients (n=19), insufficient data (n=2), case reports/series (n=3), wrong study aim (n=4), and wrong temporal relationship (n=16). The complete selection process is illustrated in the PRISMA flow diagram (Figure 1).

**Figure 1.**
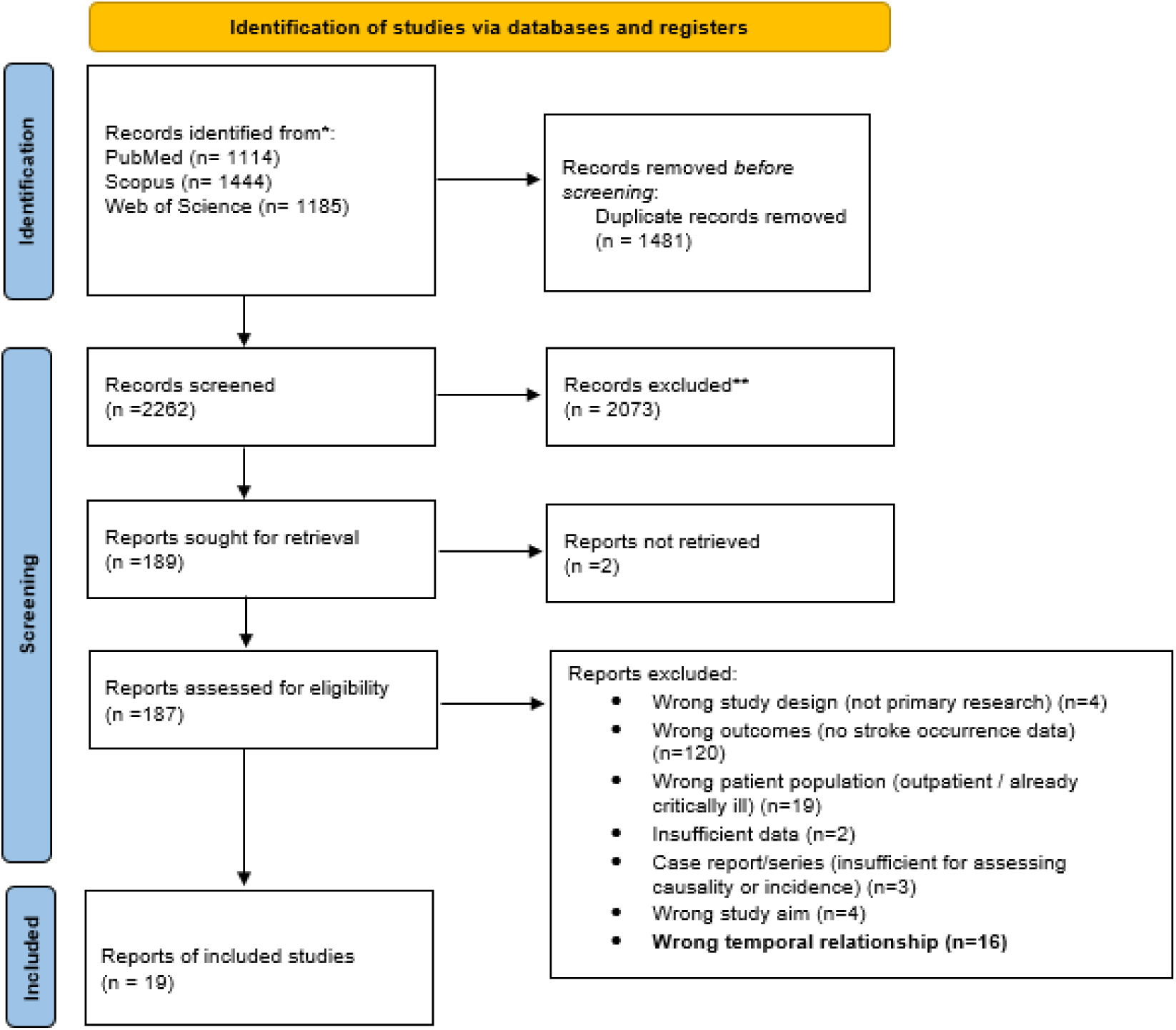
PRISMA flow chart of screening. The search strategy employed a combination of relevant terms related to COVID-19, stroke, and hospitalisation. Complete search strings for each database were constructed using appropriate controlled vocabulary and free-text terms, with adjustments made for the specific requirements and syntax of each database. The full search strings are provided in Supplementary Material Table A, with specific strings for Web of Science, PubMed and Scopus.

### Assessment of Study Quality

Population Summary: This systematic review encompassed studies published between 2020 and 2023. The studies varied considerably in size (ranging from 214 to 21,073 patients), design (predominantly retrospective cohort studies, with some prospective cohort and case-control studies), and settings (single-center, multicenter, and nationwide/international studies across Asia, Europe, and North America). Study populations included both general ward and intensive care unit patients.

Quality Assessment Tool Selection: The Newcastle-Ottawa Scale (NOS) was selected as the quality assessment tool based on its established validity for observational studies and extensive application in COVID-19 research. The NOS provides a standardised framework for evaluating methodological rigor across three domains: Selection (maximum 4 stars), which assesses representativeness of study cohorts, appropriateness of controls, ascertainment of exposure, and demonstration that outcomes were not present at baseline; Comparability (maximum 2 stars), which evaluates adjustment for confounding factors including age and cardiovascular risk factors; and Outcome (maximum 3 stars), which examines the robustness of outcome assessment, adequacy of follow-up duration, and completeness of follow-up data. This nine-point scale enables systematic comparison of study quality, with scores categorised as high (8-9 stars), moderate (6-7 stars), or low (≤5 stars) quality. The NOS has demonstrated reliability in evaluating COVID-19 studies examining cardiovascular outcomes, making it particularly suitable for our analysis of stroke occurrence. Individual study scores are presented in Supplementary Material Table S2, with detailed scoring criteria provided in Supplementary Table S3.

### Review Process: The quality assessment was conducted by a single reviewer (myself). As I was the sole reviewer, there was no need for a formal process to resolve disagreements, though this is acknowledged as a potential limitation of the review methodology

Quality Assessment Results by Research Question: As shown in Table 1, the quality assessment revealed varying levels of methodological rigor across our research questions. The strongest evidence emerged from studies examining primary stroke occurrence and comparative analyses, where most studies achieved high quality scores through clear exposure definition and adequate follow-up. Studies investigating disease severity and demographics maintained similarly high standards, while those focusing on stroke subtypes and geographical or temporal analyses showed more variable quality.

**Table 1:** Quality Assessment Table by Research Question-.

| **Research Focus** | **Number of Studies** | **High Quality (8-9★)** | **Moderate Quality (6- 7★)** | **Low Quality (≤5★)** | **Key Quality Considerations** |
| --- | --- | --- | --- | --- | --- |
| Primary Stroke Occurrence | 17 | 10 | 7 | 0 | Clear exposure definition, adequate follow-up |
| COVID-19 vs Non-COVID-19 Comparison | 3 | 1 | 2 | 0 | Appropriate hospitalised controls |
| Disease Severity (MV/ICU) | 5 | 4 | 1 | 0 | Clear severity criteria, standardised assessment |
| Demographics and Risk Factors | 6 | 4 | 2 | 0 | Complete demographic data, adjusted analyses |
| Stroke Subtypes | 7 | 5 | 2 | 0 | Imaging confirmation, standardised classification |
| Geographical/Temporal Analysis | 19 | 10 | 9 | 0 | Multi-centre representation, temporal documentation |

### Methodological Strengths and Limitations

The included studies had several methodological strengths. They featured robust exposure assessments, which were conducted through laboratory confirmation, and had clear outcome definitions that were verified with imaging. Additionally, the use of large sample sizes in multicentre studies and standardised protocols in international collaborations enhanced the quality of the evidence.

Despite these strengths, certain limitations were also present. These included variable follow-up periods and inconsistent reporting on the severity of COVID-19. Other weaknesses noted in some studies were the lack of a non-COVID-19 control group, limited adjustment for confounding factors, and potential selection bias in single-centre studies. This quality assessment demonstrates that while the included studies generally provided moderate to high-quality evidence, there was some variation in methodological rigor across different research questions. The findings suggest that future research would benefit from standardised reporting of COVID-19 severity and more comprehensive adjustment for confounding factors.

### Data Collection and Abstraction

The data abstraction process was conducted by a single reviewer (myself) using a standardised form to systematically extract relevant information from the included studies. The key data elements abstracted included study characteristics (design, period, location, sample size, and follow-up duration), population demographics (median/mean age, comorbidities, COVID-19 severity, and hospital setting), outcome measures (number of stroke events, total COVID-19 patients, control group data when available, stroke type, severity measures, and time to stroke onset), and statistical measures (effect estimates such as Odds Ratios, Hazard Ratios, and Risk Ratios, along with their 95% confidence intervals and p-values). Quality indicators, including study quality assessment scores, potential confounding factors, and adjustment methods, were also extracted. The studies predominantly consisted of retrospective cohort and observational designs conducted during 2020, with sample sizes ranging from 86 to 406,792 patients across diverse geographic locations, including the USA, Europe, Asia, and the Middle East. The extracted data encompassed populations with median/mean ages ranging from 47.2 to 78 years, with varying comorbidity profiles dominated by hypertension, diabetes, and hyperlipidaemia.

For the comparison forest plots generated in Review Manager (RevMan), sufficient raw data were extracted to calculate odds ratios (ORs) comparing stroke occurrence or incidence in hospitalised COVID-19 patients with stroke to hospitalised patients without COVID-19. This involved extracting the number of patients with and without stroke in both the COVID-19 and control groups. The same approach was used for subgroup analyses within the COVID-19 hospitalised population, where the occurrence or incidence of stroke was compared between those with and without specific risk factors (e.g., mechanical ventilation, gender, comorbidities). This involved extracting the number of COVID-19 patients with and without stroke, stratified by the presence or absence of each risk factor. For the study by Lee et al. (2023), stroke-specific data were not directly reported. Therefore, the number of stroke cases and non-stroke cases were approximated using the reported incidence rates for stroke (per 1,000,000 person-days) and the total person-days for each group, as presented in Figure 2 of the original publication. The odds ratio and 95% confidence interval for stroke were extracted directly from the published results. It is important to note that these incidence rates are adjusted using inverse probability of treatment weighting (IPTW), and therefore, the calculated stroke cases are approximations based on these adjusted rates. This potential limitation is further addressed in the Discussion section. As a single reviewer conducted the extraction, there was no need for a formal process to resolve disagreements, though this may be considered a limitation of the review methodology. Refer to the Supplementary Material Tables S4-13 for the full data extraction tables.

**Figure 2.**
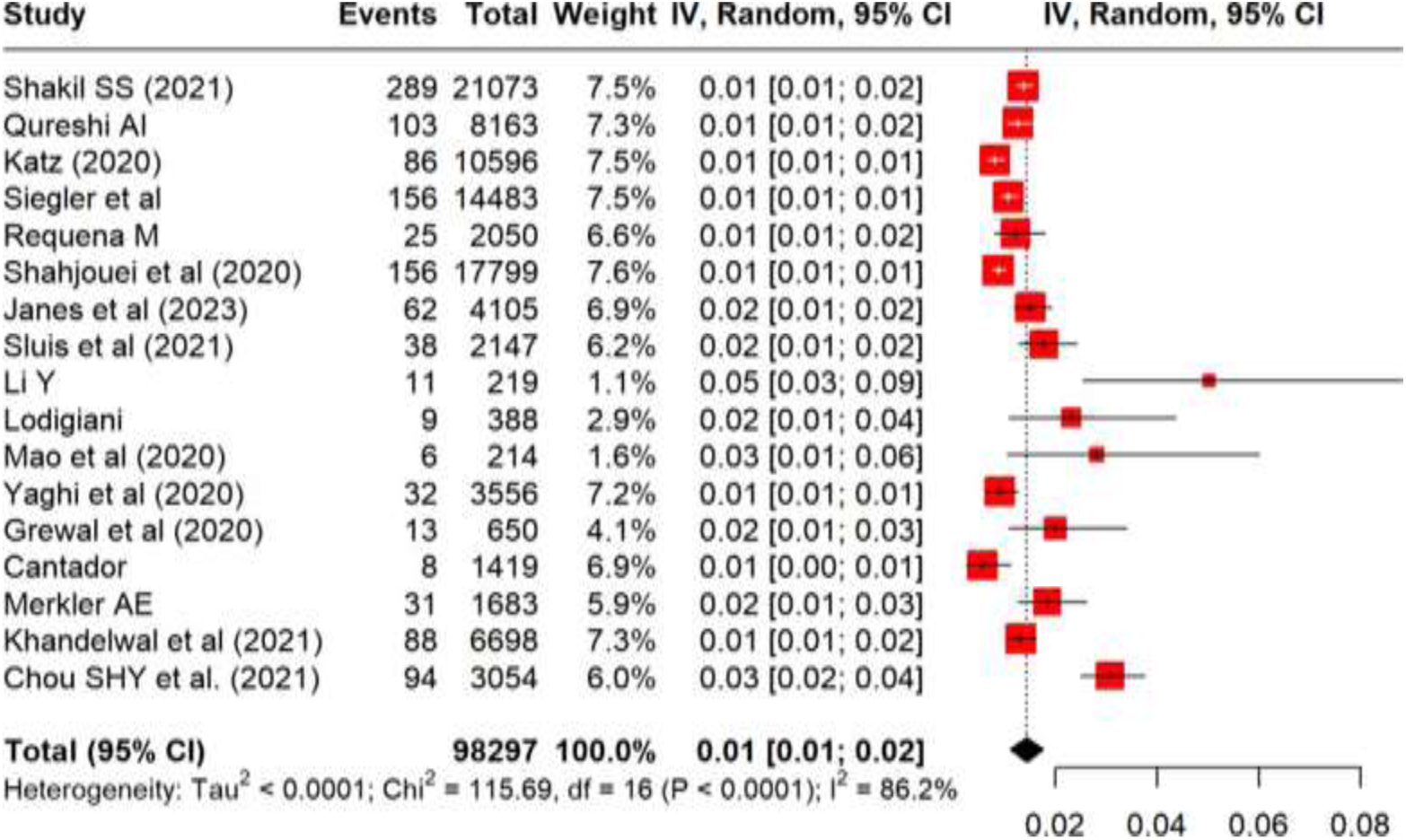
Proportion Forest Plot of Stroke Occurrence in COVID-19 hospitalised Patients. This random-effects meta-analysis summarises data from 17 studies, including a total of 98,297 patients. Each square represents the point estimate of stroke proportion from an individual study, with the size of the square being proportional to the study’s weight in the analysis. The horizontal lines extending from the squares indicate the 95% confidence intervals (CI) for each study. The diamond at the bottom represents the overall pooled proportion, which was 1% (95% CI: 1%,2%). Substantial heterogeneity was observed among the studies (I² = 86.2%, p < 0.0001).

### Meta-analysis Methodology

Statistical analyses were performed using a combination of Review Manager (RevMan) version 5.4.1 (The Cochrane Collaboration, 2020) and RStudio version 4.3.1 (R Core Team, 2023). Proportion meta-analyses were conducted using RStudio with the ’meta’ and ’metafor’ packages, while comparative analyses (odds ratios) were performed using RevMan 5.4.1.

To compare the odds of a stroke occurring between groups, the odds ratio (OR) was utilised. For the primary comparison between patients hospitalised with COVID-19 and those hospitalised without COVID-19, the odds ratio was calculated where ‘A’ represents the number of COVID-19 patients with stroke, ‘B’ is the number of COVID-19 patients without stroke, ‘C’ is the number of non-COVID-19 patients with stroke, and ‘D’ is the number of non-COVID-19 patients without stroke. For subgroup analyses comparing stroke occurrence within the COVID-19 hospitalised population, the OR was calculated similarly, but the groups were defined based on the presence or absence of a specific risk factor (e.g., mechanical ventilation, gender, comorbidity). In this scenario, ‘A’ denotes the number of COVID-19 patients with the risk factor and stroke, while ‘B’ is the number of COVID-19 patients with the risk factor but without stroke. Furthermore, ‘C’ represents the number of COVID-19 patients without the risk factor but with stroke, and ‘D’ is the number of COVID-19 patients without the risk factor and without stroke.

Statistical heterogeneity was assessed using the I² statistic and Cochran’s Q test, where I² values of 25%, 50%, and 75% were interpreted as low, moderate, and high heterogeneity, respectively. The τ² (tau-squared) statistic was used to quantify the between-study variance in effect sizes. Statistical significance for heterogeneity was set at p<0.05 for the Q test. To account for anticipated clinical and methodological diversity across studies, a random-effects model using the DerSimonian and Laird method was employed for all analyses. Pre-planned sensitivity analyses were conducted to explore potential sources of heterogeneity across three dimensions: geographical region (North America, Europe, Asia, and multi-national studies), pandemic phase (2020, 2021, and 2022), and study type (multi-centre, registry-based, and single-centre studies). Publication bias was assessed through visual inspection of funnel plots for all major analyses. Effect estimates were presented as pooled proportions with 95% confidence intervals (CIs) for the primary analysis, and odds ratios (ORs) with 95% CIs for comparative analyses. Statistical significance was set at p<0.05.

## Results

The literature search that was conducted between September 2024 to January 2025 identified 3,743 publications, including 1,114 from PubMed, 1,444 from Scopus, and 1,185 from Web of Science. After removing 1,481 duplicate records, 2,262 records remained for screening. Of these, 2,073 were excluded based on initial screening, leaving 189 reports for retrieval. Two reports could not be retrieved, resulting in 187 full-text articles assessed for eligibility. Following full-text review, 170 articles were excluded for the following reasons: wrong study design (n=3), no stroke occurrence data (n=98), wrong patient population (n=32), insufficient data (n=12), case reports/series insufficient for assessing causality (n=11), wrong study aim (n=6), and wrong temporal relationship (n=8). Therefore, 19 studies { (14), (15),(10), (16),(17), (18) (19) (20) (21) (12) (4) (11) (22) (23) (5), (24), (25), (26), (27) } were included overall in the final systematic review and meta-analysis, 17 were included in the proportion forest plot and 2 studies {(26), (27)} (1 from the primary proportion forest plot batch) where included in a preliminary analysis.

### Characteristics of Included Studies

Overall, 17 studies with a total of 97,743 hospitalised COVID-19 patients were included in this meta-analysis. The sample sizes of individual studies ranged considerably, from 214 to 21,073 patients. All these studies were conducted between 2020 and 2023, with nine studies published in 2020-2021 and two studies published in 2022-2023. The largest studies were conducted by Shakil SS (14) (2021; n=21,073), Shahjouei et al (18) (2020; n=17,799), and Siegler et al (16) (n=14,483).

Patient characteristics are presented in the Supplementary Materials Tables, detailed characteristics of included studies are presented in Supplementary Material Table S4. Complete subgroup analyses including mechanical ventilation status, gender distribution, ICU admission rates, age groups, and comorbidities, along with their respective raw data are presented in Figures 4-7 and Supplementary Materials.

### Baseline stroke occurrence rates

Seventeen studies evaluated stroke occurrence in patients hospitalised with COVID-19, with rates varying between 0.6% and 5%. The combined analysis included 98,297 patients, with stroke documented in 1,211 cases, resulting in a pooled occurrence rate of 1% (95% CI: 1%-2%, p<0.001), shown in Figure 2. The included research spanned multiple nations and diverse patient populations. Substantial statistical heterogeneity was observed between studies (I² = 86.2%, p<0.0001), indicating considerable variation in stroke risk depending on patient and clinical factors. Notable larger cohorts, such as Shakil SS (14) (289/21,073, 1.4%), Shahjouei et al (18) (156/17,799, 0.9%), and Siegler et al (16) (156/14,483, 1.1%), showed similar stroke frequencies approximately 1%. The stroke rate from Chou’s (25) all COVID-19 cohort was 3% (94/3,054). Further analyses examining geographical distribution, time of publication, sample size, and methodology confirmed the consistency of these primary results. Patient characteristics and detailed subgroup analyses are presented in the Supplementary Materials. Assessment of statistical heterogeneity revealed substantial variation in this primary analysis (I² = 86.2%, p<0.0001), with individual study rates ranging from 0.6% to 5.0%. Visual inspection of the funnel plot for this analysis suggested some asymmetry, with smaller studies reporting higher stroke occurrence rates.

### COVID-19 hospitalisation and risk of stroke compared

COVID-19 hospitalisation was not significantly associated with an increased risk of stroke compared to non-COVID-19 hospitalised controls, though the evidence showed substantial heterogeneity. Meta-analysis of three studies comprising 1,107 patients yielded a pooled odds ratio of 1.07 (95% CI: 0.41,2.81, p = 0.89), indicating no clear difference in stroke occurrence between groups. Individual study results varied considerably (I² = 90%, p < 0.0001): Merkler et al. (5) reported significantly higher stroke risk in COVID-19 patients (OR = 8.13, 95% CI: 2.48,26.60), while more recent studies by Ramos et al. (2021) (26) and Lee et al. (27) found lower stroke risk (OR = 0.49, 95% CI: 0.32,0.76 and OR = 0.60, 95% CI: 0.42,0.86, respectively). The temporal variation in findings, with earlier pandemic studies showing higher risk than later ones, suggests potential changes in COVID-19 management over time may have influenced stroke risk. Study weights were relatively balanced, with Lee et al. (27) contributing 38.0%, Ramos et al. (26) 37.0%, and Merkler et al. (5) 25.0% to the overall analysis. High heterogeneity was observed in this comparative analysis (I² = 90%, p<0.0001), likely reflecting temporal evolution in COVID-19 management protocols across the pandemic period. Funnel plot analysis showed relatively symmetric distribution of effect sizes, suggesting limited publication bias for this comparison. COVID-19 Hospitalised VS Non-COVID-19

The meta-analysis comparing stroke risk between COVID-19 hospitalised patients and non-COVID-19 controls revealed no significant overall difference, with a pooled odds ratio of 1.13 (95% CI: 0.49,2.65, p = 0.77), shown in Figure 3. However, there was substantial heterogeneity among the included studies (I² = 89%, p < 0.0001), suggesting considerable variation in findings across different clinical settings and time periods.

**Figure 3.**
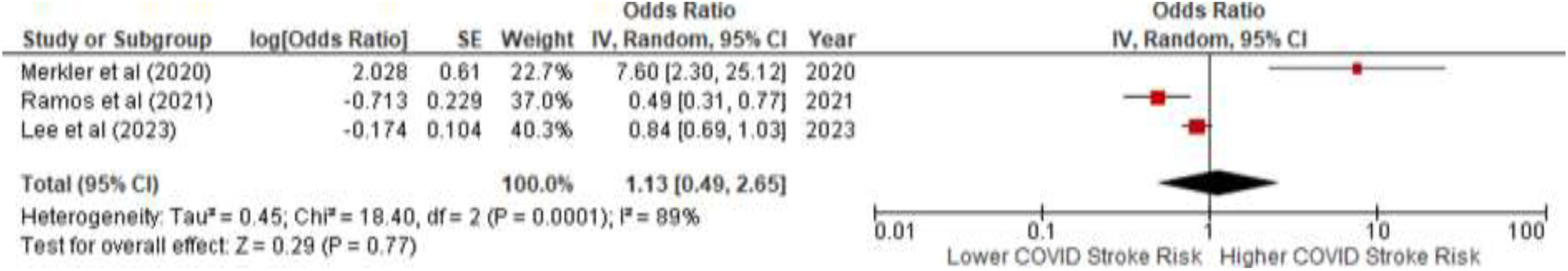
Association Between COVID-19 Hospitalisation and Stroke Risk Compared to Non-COVID-19 Hospitalised Controls. A Meta-Analysis of Three Studies. This random-effects meta-analysis includes three studies. Individual squares represent the odds ratio (OR) from each study, with the size of the square proportional to the study’s weight in the analysis. The horizontal lines indicate the 95% confidence intervals (CI). The diamond shows the pooled OR of 1.13 (95% CI: 0.49,2.65), indicating no significant overall difference in stroke risk between the groups. Substantial heterogeneity was observed among the studies (I² = 89%, p < 0.0001).

Individual study results varied markedly over time. The earliest study by Merkler et al. (5) reported significantly higher stroke risk in COVID-19 patients (OR = 7.60, 95% CI: 2.30,25.12), while more recent studies found lower risks. Ramos et al. (26) reported reduced stroke risk (OR = 0.49, 95% CI: 0.31,0.77), and Lee et al. (27) found a slight but non-significant reduction (OR = 0.84, 95% CI: 0.69,1.03).

The analysis weighted the studies based on their precision, with Lee et al. contributing 40.3%, Ramos et al. 37.0%, and Merkler et al. 22.7% to the overall estimate. The temporal pattern of decreasing risk estimates from 2020 to 2023 suggests possible improvements in COVID-19 management protocols over time. However, the high heterogeneity (Tau² = 0.45, Chi² = 18.40, df = 2, p < 0.0001) indicates that these findings should be interpreted with caution and that other factors may influence the relationship between COVID-19 hospitalisation and stroke risk.

### Stroke risk factors among COVID-19 hospitalised population

The occurrence was assessed through comparative analyses of subgroups (ICU admission, mechanical ventilation, gender, and comorbidities).

### Mechanical Ventilation

COVID-19 patients who experienced stroke during hospitalisation were significantly more likely to require mechanical ventilation compared to those who did not develop stroke. Meta-analysis of four studies demonstrated that patients requiring mechanical ventilation had 3.59 times higher odds of developing stroke compared to non-ventilated patients (OR: 3.59, 95% CI: 2.21,5.83, p<0.00001), shown in Figure 4. The studies showed consistent direction of effect, with odds ratios ranging from 2.02 (Qureshi et al. (15)) to 5.95 (Shakil et al. (14)).

**Figure 4.**
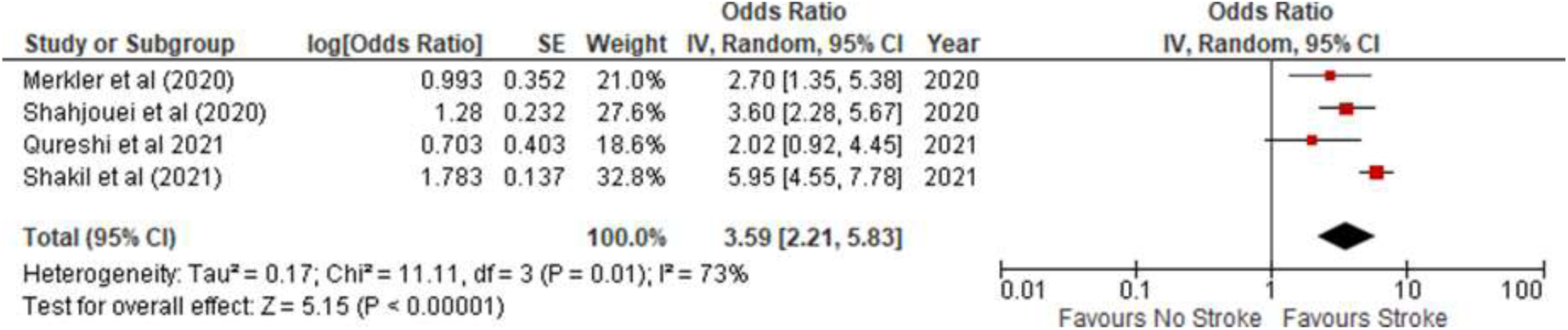
Association Between Mechanical Ventilation and Stroke Risk Among Hospitalised COVID-19 Patients. There was substantial heterogeneity among studies (I² = 73%, p=0.01), suggesting variability in the strength of this association across different clinical settings and populations. Despite this heterogeneity, the consistent direction of effect across all studies and the highly significant overall effect (Z = 5.15, p<0.00001) strongly support mechanical ventilation as an important marker of stroke risk in COVID-19 patients. The log odds ratios ranged from 0.703 to 1.783, indicating a consistently positive association between mechanical ventilation and stroke risk across all studies.

All studies contributed meaningful weight to the analysis, with Shakil et al. (14) contributing the highest weight at 32.8%, followed by Shahjouei et al. (18) at 27.6%, Merkler et al. (5) at 21.0%, and Qureshi et al. (15) at 18.6%. The largest study by Shakil et al. (14) reported the highest odds ratio of 5.95 (95% CI: 4.55,7.78), while Shahjouei et al. (18) found an OR of 3.60 (95% CI: 2.28,5.67).

### ICU Admission

COVID-19 patients who experienced stroke during hospitalisation were substantially more likely to require ICU admission compared to those who did not develop stroke. Meta-analysis of three studies (n=23,224 total patients) demonstrated that those admitted to the ICU had 6.33 times higher odds of developing stroke compared to non-ICU patients (OR: 6.33, 95% CI: 4.95,8.09, p<0.00001), shown in Figure 5. The largest study by Shakil et al. (14), which contributed 85.6% of the weighted analysis, showed that among stroke patients, 74.4% (110/160) required ICU admission compared to 30.2% (6,270/20,784) of non-stroke patients. This finding was supported by Merkler et al. (5), who reported 61% (19/31) ICU admission in stroke patients versus 24% (455/1,885) in non-stroke patients, and Lodigiani et al. (12), who found similar trends. The association between ICU admission and stroke was highly consistent across studies, with minimal heterogeneity observed (I²= 0%, p=0.38), supporting the robustness of this finding despite differences in study settings and populations.

**Figure 5.**
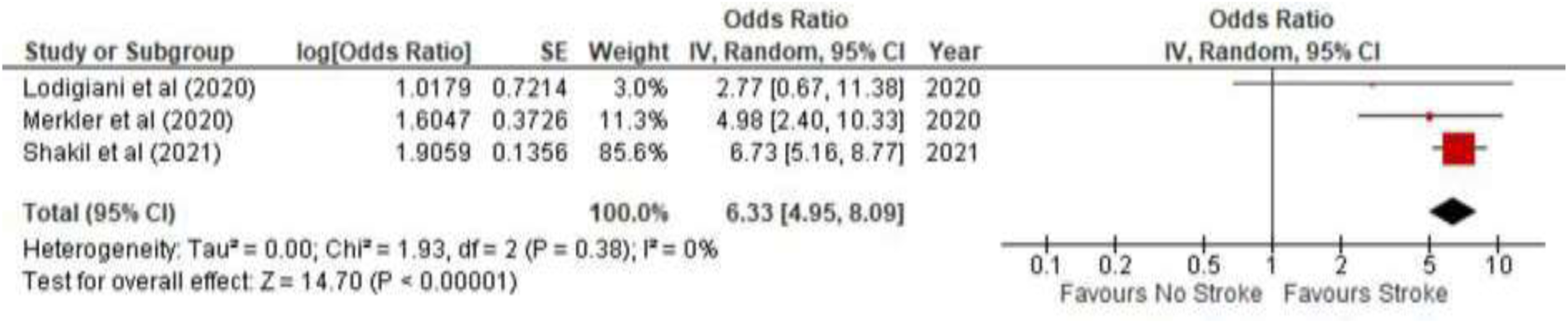
Forest Plot of the Association Between ICU Admission and Stroke Occurrence in Hospitalised COVID-19 Patients. The figure presents a meta-analysis of three studies comparing the odds of requiring intensive care unit (ICU) admission between hospitalised COVID-19 patients who developed a stroke and those who did not. Individual study odds ratios (ORs) and their 95% confidence intervals (CIs) are depicted by squares and horizontal lines, respectively. The size of each square is proportional to the study’s weight in the analysis. The diamond represents the pooled OR calculated using a random-effects model, with its width indicating the overall 95% CI. The analysis shows that patients who developed a stroke had significantly higher odds of being admitted to the ICU (pooled OR: 6.33; 95% CI: 4.95,8.09; *P* < 0.00001). No significant heterogeneity was observed among the studies (I² = 0%, *P* = 0.38).

### Gender Sub-group Analysis

To investigate the influence of gender on stroke occurrence among hospitalised COVID-19 patients, a meta-analysis of six studies comprising 49,620 individuals (26,519 males and 23,101 females) was conducted. The analysis revealed no statistically significant difference in the odds of stroke between male and female patients (pooled OR: 1.17, 95% CI: 0.87,1.57; *P*=0.29), as shown in Figure 6. Although the overall association was not significant, the included studies demonstrated notable variability in the direction and magnitude of the gender effect. Two large, early pandemic studies by Shahjouei et al. (18) and Shakil et al. (14) reported a significantly higher odds of stroke in males, with ORs of 1.69 (95% CI: 1.20,2.38) and 1.46 (95% CI: 1.06,2.02), respectively. Conversely, more recent studies by Sluis et al. (20) and Janes et al. (19) suggested a non-significant trend toward a higher stroke occurrence in females (OR: 0.76, 95% CI: 0.40,1.46 and OR: 0.81, 95% CI: 0.44,1.47, respectively).

**Figure 6.**
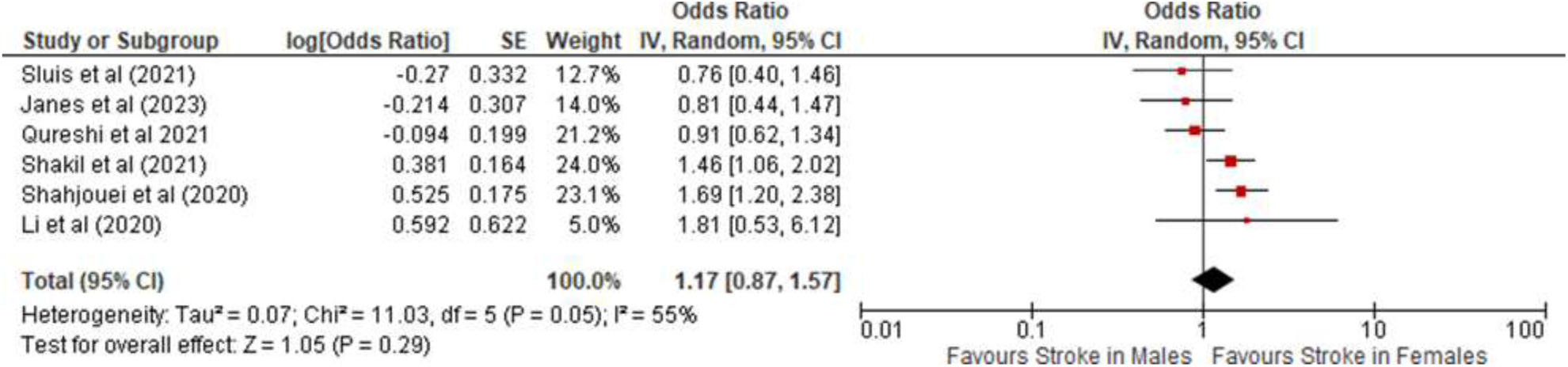
Association Between Gender and Stroke Risk in COVID-19 Patients. The meta-analysis of six studies compares the odds of stroke in male versus female patients. Individual study odds ratios (ORs) are represented by squares, with the size of each square proportional to the study’s weight in the analysis. The horizontal lines indicate the 95% confidence intervals (CIs). The diamond represents the pooled OR from a random-effects model, with its width corresponding to the 95% CI. The overall analysis of 49,620 patients found no significant difference in stroke occurrence between genders (pooled OR: 1.17; 95% CI: 0.87,1.57; *P*=0.29), with moderate heterogeneity across studies (I² = 55%; *P*=0.05).

Moderate heterogeneity was observed across the included studies (I² = 55%, *P*=0.05), suggesting that the relationship between gender and stroke risk may be influenced by other factors, such as the clinical setting, patient population, or viral variant. In total, 479 stroke events were recorded across the six studies, with 290 cases occurring in males and 189 in females.

### Comorbidities Sub-group Analysis

The meta-analysis examining comorbidities across four studies reveals important insights into the relationship between pre-existing conditions and stroke risk in hospitalised COVID-19 patients, shown in Figure 7. Overall, the analysis demonstrates a significant increase in stroke odds among patients with comorbidities, with a pooled odds ratio of 1.75 (95% CI: 1.31,2.34, p=0.0002), though there was substantial heterogeneity in the findings (I² = 82%).

**Figure 7.**
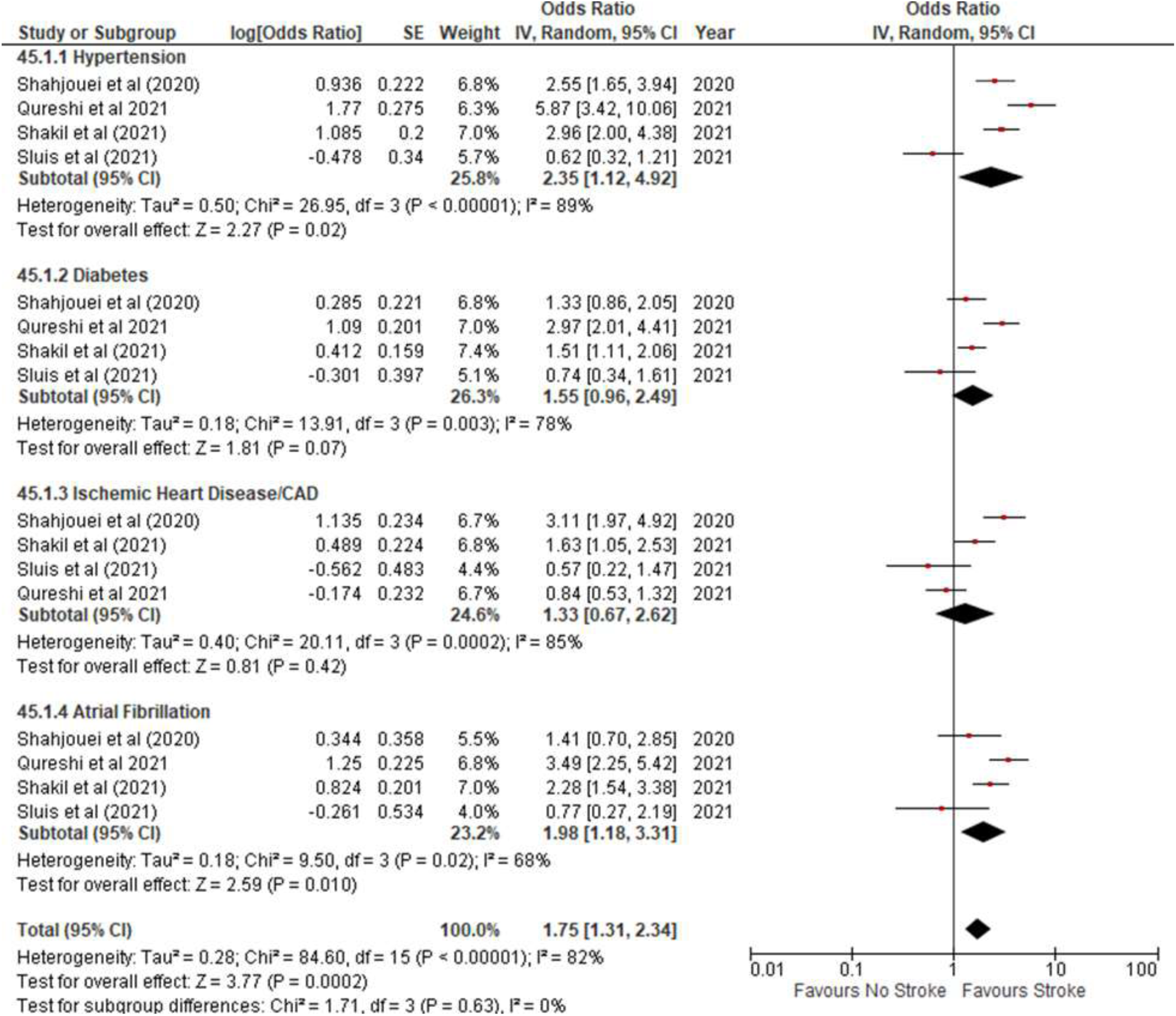
Forest Plot of the Association Between Pre-existing Comorbidities and Stroke Risk in Hospitalised COVID-19 Patients. The subgroup meta-analysis examines the association between four common pre-existing comorbidities (hypertension, diabetes, ischemic heart disease/coronary artery disease, and atrial fibrillation) and the odds of stroke. Individual study odds ratios (ORs) are represented by squares, with horizontal lines indicating the 95% confidence intervals (CIs). The diamonds represent the pooled OR for each subgroup and for the overall analysis, calculated using a random-effects model. Overall, the presence of any comorbidity was significantly associated with increased odds of stroke (pooled OR: 1.75; 95% CI: 1.31,2.34; *P*=0.0002). Subgroup analyses identified hypertension (OR: 2.35; 95% CI: 1.12,4.92) and atrial fibrillation (OR: 1.98; 95% CI: 1.18,3.31) as significant risk factors. The analysis showed high overall heterogeneity (I² = 82%), but the test for subgroup differences was not significant (*P*=0.63). Abbreviations: CAD, coronary artery disease; CI, confidence interval; OR, odds ratio.

Among the studied conditions, hypertension emerged as a significant risk factor, showing the strongest association with stroke occurrence (OR: 2.35, 95% CI: 1.12,4.92, p=0.02). The impact of hypertension varied considerably across studies, with Qureshi et al. (15) reporting the highest odds ratio of 5.87.

Diabetes showed a trend toward increased stroke risk, though the association just missed statistical significance (OR: 1.55, 95% CI: 0.96,2.49, p=0.07). The relationship between diabetes and stroke risk was most pronounced in the Qureshi et al. (15) study, which reported an odds ratio of 2.97.

For Ischemic Heart Disease/CAD, the analysis found no significant association with stroke occurrence (OR: 1.33, 95% CI: 0.67,2.62, p=0.42). This finding showed considerable variation across studies, contributing to the high heterogeneity observed.

Atrial fibrillation demonstrated a significant association with stroke risk (OR: 1.98, 95% CI: 1.18,3.31, p=0.010), with Qureshi et al. (15) again reporting the strongest effect (OR: 3.49). The heterogeneity for this comorbidity was moderate (I² = 68%). Importantly, the test for subgroup differences was not significant (Chi² = 1.71, p=0.63, I²= 0%), suggesting that the strength of association was relatively consistent across different types of comorbidities. The consistent presence of heterogeneity across all subgroups underscores the complex nature of how these risk factors interact with stroke occurrence in COVID-19 patients. Publication bias was assessed using Egger’s test and visual inspection of the funnel plot, which can be found in Supplementary Materials (Supplementary Figure S12).

Additional analyses were conducted to examine stroke subtypes, age distribution, and potential sources of heterogeneity. Ischemic stroke was the predominant subtype with a pooled proportion of 81% (95% CI: 73,90%), followed by haemorrhagic stroke (20%, 95% CI: 12,28%), while CVST showed the lowest occurrence (2%, 95% CI: 0,3%). Age distribution analysis revealed a clear age-dependent pattern, with the lowest stroke occurrence in patients <35 years (2%, 95% CI: 0,7%), increasing progressively through age groups 35-54 (18%, 95% CI: 10,28%) and 55-70 (40%, 95% CI: 30,51%), before slightly decreasing in patients >70 years (39%, 95% CI: 29,48%). Sensitivity analyses by geographical region showed consistent stroke rates across North America (1%, 95% CI: 1,2%), Europe (1%, 95% CI: 1,2%), and multi-national studies (2%, 95% CI: 1,3%), with slightly higher rates in Asian studies (4%, 95% CI: 0,49%), though with wider confidence intervals due to smaller sample sizes. Significant heterogeneity was observed across studies (I² = 90.0%, p < 0.0001). Detailed forest plot figures, figure legends and complete analyses are available in Supplementary Materials Figure S1-7. Sensitivity Analyses Further investigation of heterogeneity sources revealed systematic variations across study characteristics. Multi-centre studies demonstrated the highest heterogeneity (I² = 96.8%), followed by registry-based studies (I² = 87.2%), and single-centre studies (I² = 79.1%). Geographical analysis revealed greater heterogeneity in North American studies compared to European and Asian studies. Temporal analysis showed decreasing heterogeneity from 2020 (I² = 72%) to 2022 (I² = 0%), suggesting standardisation of reporting practices over time.

Publication Bias Visual inspection of funnel plots across all analyses suggested potential publication bias, particularly for smaller studies reporting higher stroke occurrence rates.

For the primary analysis of stroke occurrence, the funnel plot reveals moderate asymmetry around the pooled proportion estimate of 1% Figure 8. While larger studies with smaller standard errors cluster near the pooled estimate, smaller studies show greater dispersion across a wider range of proportions (1–5%). However, this observed asymmetry may reflect not only potential publication bias but also true clinical heterogeneity in the relationship between COVID-19 hospitalisation and stroke occurrence across different clinical settings and populations.

**Figure 8.**
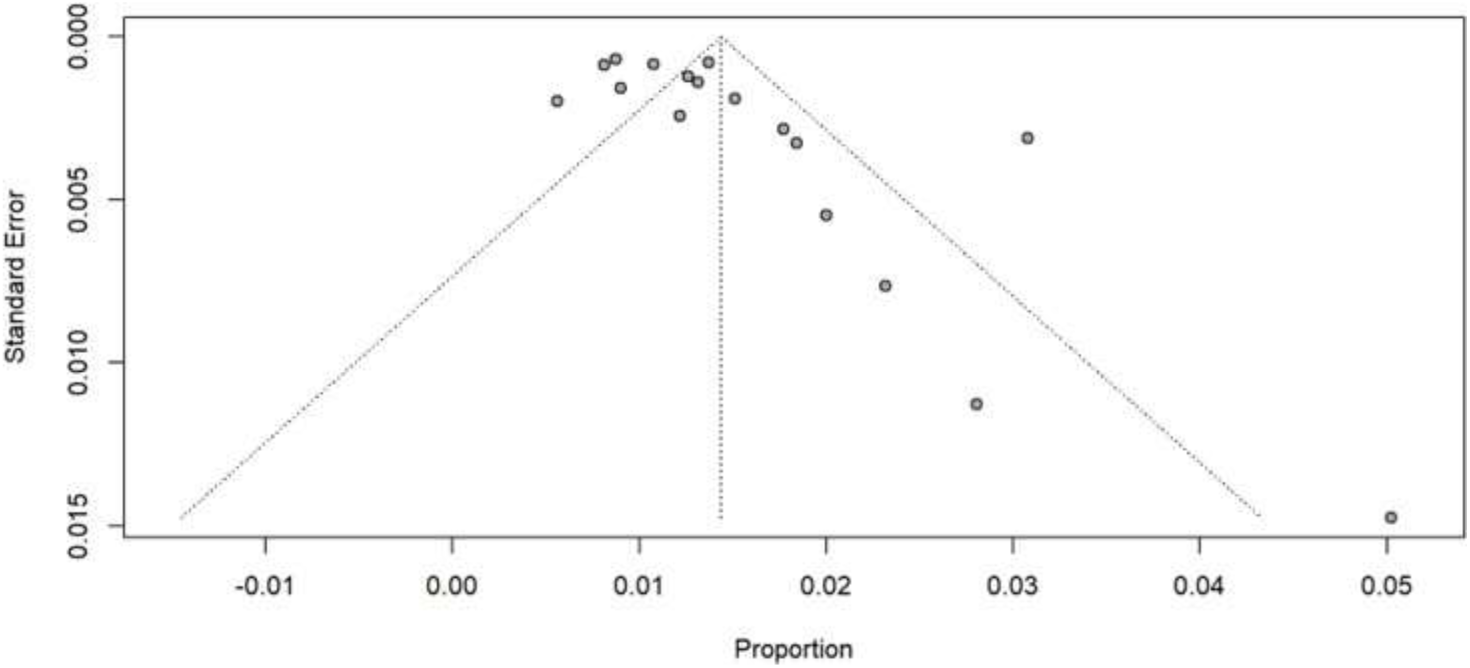
Publication Bias Assessment for Primary Forest plot. Visual inspection of the funnel plot reveals moderate asymmetry around the pooled proportion estimate of 0.01 (1%). While larger studies (smaller standard errors) cluster consistently around the pooled estimate, smaller studies show greater dispersion across proportion ranges (0.01-0.05). This pattern, combined with the substantial statistical heterogeneity (I² = 86.2%, p <0.0001), suggests the asymmetry may reflect both potential publication bias and true clinical heterogeneity across different study populations and settings.

Additional publication bias assessments are provided in the Supplementary Material Figures S8-12, which includes funnel plots for the COVID-19 versus non-COVID-19 comparison, mechanical ventilation analysis, ICU admission analysis, gender analysis, and comorbidity analysis.

## Discussion

This systematic review and meta-analysis investigated the intricate relationship between COVID-19 hospitalisation and stroke occurrence, aiming to clarify the risk and identify contributing factors. The primary analysis, encompassing 98,297 patients across 17 studies, revealed a pooled stroke occurrence rate of 1% among hospitalised COVID-19 patients. This finding is validated by Shahjouei et al.’s (28) multinational study of 26,175 patients across 99 tertiary hospital centres, which demonstrated a comparable stroke risk of 0.5-0.9%. The initially reported higher rates have been refined through larger cohorts, with Li et al.’s (29) early report of 4.6% being contextualised by subsequent studies showing consistently lower rates: Yaghi et al. (11) found 0.9% risk of stroke among 3,556 patients, and Merkler et al. (5) reported 1.6% risk of stroke among 1,916 patients.

### Baseline Stroke Occurrence Rates in COVID-19 Hospitalisation

Our meta-analysis reveals a 1% pooled stroke rate during COVID-19 hospitalisation, representing a middle ground among varying reports. Early studies (Li et al. (29); Mao et al. (4)) showed higher rates due to selection bias and limited testing. Later research provided different perspectives: Shakil et al.’s (14) registry study (n=21,073) found 0.75% occurrence, while Simonsen et al.’s (30) nationwide analysis showed stable admission rates despite increased mortality. Uchino et al.’s (31) observed a 30% decrease in stroke presentations during the pandemic surge, indicating changed healthcare-seeking behaviour. Systematic reviews varied significantly - Tan et al.’s (2020) reported 1.4% versus Nagraj et al.’s (32) 0.175% in RCTs. Klok et al.’s (2) finding of 31% thrombotic complications in ICU patients suggests severity-dependent relationships.

Comparing Shakil et al.’s (14) US registry study across 107 hospitals with Simonsen et al.’s (30) Danish nationwide study reveals healthcare system influences on stroke occurrence. While Shakil reported 0.75% ischemic stroke/TIA rate, Simonsen found no significant change in admission rates. Both studies challenged age-related assumptions: Shakil identified higher risk in middle-aged adults, while Simonsen noted increased mortality during initial lockdowns.

Our study’s substantial heterogeneity highlights methodological challenges. Qureshi et al. (15) identified specific cardiovascular risk factors, while Klok et al.’s (2) observation of 31% thrombotic complications in ICU patients indicates severity-dependent relationships. Li et al.’s (29) found significantly older age and more severe disease in COVID-19 stroke patients (81.8% vs 39.9%, p<0.01). Uchino et al.’s (31) documentation of unchanged stroke severity despite reduced presentations suggests selection bias toward severe cases.

Critical questions remain regarding COVID-19 hospitalisation and stroke occurrence. While our meta-analysis indicates a 1% pooled rate, true stroke risk remains uncertain, particularly given missed mild cases. The pathophysiological mechanisms require clarification, despite markers like elevated D-dimer levels (mean 9.2±14.8 mg/L) suggesting unique pathways. Long-term stroke risk following COVID-19 hospitalisation needs further study. Future research should prioritise prospective studies with robust surveillance systems, systematic monitoring of inflammatory and coagulation markers, and extended follow-up periods. Developing specific risk stratification tools and investigating vaccination status’s impact on stroke risk during hospitalisation are crucial next steps.

### Stroke Risk in COVID-19 vs. non-COVID-19 Hospitalisations

Our meta-analysis challenges initial pandemic assumptions, showing no significant difference in stroke risk between COVID-19 and non-COVID-19 hospitalised patients. The substantial heterogeneity reflects varying study methodologies and evolving understanding of COVID-19-related thrombotic risk.

Early pandemic research by Merkler et al. (5) reported significantly elevated stroke risk in COVID-19 patients compared to influenza cases (OR 7.6, 95% CI: 2.3,25.2). However, Lo Re III et al.’s (33) subsequent large-scale study of 85,637 COVID-19 patients found no significant difference in arterial thromboembolism risk (aHR 1.04-1.07) versus influenza patients. While this larger study employed improved methodology, it combined stroke with myocardial infarction in a composite outcome.

Ramos et al. (26) documented 1.5% stroke incidence with distinct characteristics, including higher stroke severity (elevated NIHSS scores, p < 0.001) and increased hemorrhagic conversion (p = 0.034). Lee et al.’s (27) nationwide study found no increased overall cardiovascular risk (HR 0.84, 95% CI: 0.69,1.03), supporting recent evidence that initial risk estimates were influenced by early pandemic circumstances.

Clinical characteristics show distinct patterns, with Akhtar et al. (34) noting more severe presentations (NIHSS >10: 34.4% vs 16.7%) and poorer outcomes (mRS 0-2: 28.1% vs. 51.9%). Regarding pathophysiology, Lodigiani et al. (12) observed early thromboembolic events (50% within 24 hours), while Beyrouti et al.’s (3) findings of multiple territory infarcts suggested unique COVID-19 mechanisms. Dengler et al.’s (35) observation of declining concurrent COVID-19-stroke cases (from 3.4% to 0.4%) indicates successful adaptation of preventive strategies.

Critical knowledge gaps and research priorities encompass methodological needs, including the necessity for large-scale studies with stroke-specific outcomes, standardisation of outcome measures, investigation of vaccination impact, and longer-term follow-up studies, alongside clinical priorities such as developing risk-stratified prevention strategies, optimising antithrombotic protocols, and investigating mechanism-specific interventions.

While early pandemic data from Merkler et al. (5) suggested dramatically increased risk (OR 7.6, 95% CI: 2.3,25.2), our comprehensive analysis reveals a more nuanced picture. The evolution to more robust methodologies, demonstrated by Lo Re III et al.’s (33) large-scale study, suggests COVID-19 hospitalisation may not substantially increase stroke risk compared to other severe respiratory infections. However, the use of composite arterial endpoints challenges isolation of stroke-specific risk.

Our analysis shows that while COVID-19 hospitalisation may not independently increase stroke occurrence, it presents unique characteristics requiring specific attention. The progression from early single-centre studies to recent large-scale analyses has refined our understanding, though widespread use of composite endpoints limits definitive conclusions. Future research must prioritise stroke-specific outcomes while maintaining robust methodological standards to enable targeted prevention strategies based on individual risk profiles rather than COVID-19 status alone.

### Factors Influencing Stroke Occurrence in Hospitalised COVID-19 Patients

Our meta-analysis provides compelling evidence regarding factors that influence stroke occurrence during COVID-19 hospitalisation, with mechanical ventilation emerging as a crucial predictor. The analysis of five studies demonstrated that mechanically ventilated COVID-19 patients had significantly higher odds of stroke. This finding is supported across varying study sizes, from Shahjouei et al.’s (28) large multinational study of 26,175 patients (OR: 1.9, 95% CI:1.1,3.5) to smaller detailed clinical studies, though reported stroke incidence varies notably (0.9-3.4%) across different healthcare settings.

The pathophysiological mechanisms linking mechanical ventilation and stroke in COVID-19 patients appear multifaceted and complex. While Fan et al. (36) suggested a prothrombotic mechanism through antiphospholipid antibodies in ventilated patients (83.3% vs. 26.9%), Topcuoglu et al.’s (37) prospective case-control study found no significant increase in antiphospholipid antibodies, suggesting the relationship between mechanical ventilation and stroke may reflect disease severity rather than a unique COVID-19-specific mechanism. This interpretation is supported by Li et al.’s (29) observation of elevated inflammatory markers in ventilated patients who developed stroke (CRP: 51.1 vs 12.1 mg/L, p<0.05; D-dimer: 6.9 vs 0.5 mg/L, p<0.001).

The relationship between prior stroke history, mechanical ventilation, and comorbidities presents a complex interplay requiring careful interpretation. While our analysis identified hypertension as the strongest predictor, Qin et al.’s (38) study of COVID-19 patients with prior stroke history suggests this may reflect pre-existing cerebrovascular disease rather than a direct COVID-19 effect. Topcuoglu et al.’s finding of similar vascular risk factors between COVID-19 positive and negative stroke patients further supports the importance of traditional risk factors. This is reinforced by Jamora et al.’s (39) observation that prior stroke history (OR: 2.3, 95% CI: 1.82,2.97) carried a higher risk than hypertension (OR: 1.75, 95% CI: 1.53,1.97) for stroke occurrence.

Our findings address several key knowledge gaps in the literature, though important limitations remain. While our meta-analysis provides robust evidence of mechanical ventilation’s predictive value for risk of stroke, the heterogeneity in ventilation protocols across studies suggests caution in interpretation. The relative importance of traditional versus COVID-19-specific risk factors remains unclear, particularly given Topcuoglu et al.’s evidence suggesting no unique COVID-19 stroke mechanisms.

These findings have immediate public health implications, though implementation challenges must be considered. While enhanced monitoring of ventilated COVID-19 patients appears warranted, the similar risk factor profiles between COVID-19 and non-COVID-19 stroke patients suggest that traditional stroke prevention strategies remain relevant. Healthcare systems must balance resource allocation between COVID-19-specific monitoring and established stroke prevention protocols.

This analysis advances our understanding of stroke risk in hospitalised COVID-19 patients while highlighting critical knowledge gaps. Future research must address several key limitations through prospective studies with standardised protocols, particularly focusing on distinguishing between the effects of mechanical ventilation, disease severity, and traditional risk factors on stroke occurrence in COVID-19 patients.

### Age and Gender Distribution of Stroke Occurrence (Demographics)

The demographic patterns of stroke occurrence in COVID-19 hospitalised patients reveal complex distributions challenging traditional stroke risk paradigms. Our meta-analysis showed no significant overall gender difference in stroke occurrence, though temporal and variant-specific patterns emerged throughout the pandemic.

Early pandemic studies (2020) demonstrated male predominance in stroke occurrence (Shahjouei et al.: OR 1.69, 95% CI: 1.20,2.38; Li et al.: OR 1.81, 95% CI: 0.53,6.12; Shakil et al.: OR 1.46, 95% CI: 1.06,2.02). This pattern shifted in later studies (2021-2023) toward female predominance or neutrality (Sluis et al.: OR 0.76, 95% CI: 0.40, 1.46; Janes et al.: OR 0.81, 95% CI: 0.44,1.47; Qureshi et al.: OR 0.91, 95% CI: 0.62,1.34). Kopp et al.’s (40) analysis showed variant-specific gender differences, with higher male cardiovascular mortality during Delta (6.13% vs 3.62% in women, p=0.017), but no significant gender differences during Alpha and Omicron variants.

Age-stratified analysis revealed peak stroke rates in the 55-70 age group (40%, 95% CI: 30,51%), followed by those >70 years (39%, 95% CI: 29,48%), and lower rates in younger groups (<35 years: 2%, 95% CI: 0,7%). This aligns with Merkler et al.’s (5) observation of younger stroke presentations (median age 69 years, IQR: 66-78). Li et al. (29) found COVID-19 stroke patients were significantly older (75.7 ± 10.8 years) than non-stroke COVID-19 patients (52.1 ± 15.3 years, p<0.001).

Wilcox et al.’s (41) findings showed gender differences were most pronounced in younger age groups (aged 18-54 years: aOR 3.17, 95% CI 2.06,5.01) but attenuated in older populations (aged ≥75 years: aOR 1.20, 95% CI 0.90,1.59). Mao et al. (4) observed acute cerebrovascular disease occurred in 5.7% of severe COVID-19 cases versus 0.8% in non-severe cases.

High study heterogeneity (I² 78.7-86%) reflects viral variant evolution, improved clinical recognition, and evolving treatment protocols. Clinical protocols must consider viral variants and demographic factors. Prevention strategies need demographic tailoring and variant adaptability. Research priorities include studying variant pathophysiology and long-term outcomes across populations. Treatment responses require demographic-specific evaluation. COVID-19’s stroke risk impact varies by individual characteristics and viral variants, demanding personalised assessment approaches.

### Distribution and Contribution of Stroke Subtypes

Our meta-analysis of 710 COVID-19 patients reveals ischemic stroke as the predominant subtype, with a pooled prevalence of 81% (95% CI: 0.73,0.90), showing significant heterogeneity (I² = 89.4%, p < 0.0001) across healthcare settings. While Tan et al.’s (13) comprehensive meta-analysis reported 87.4% ischemic strokes, Shakil et al.’s (14) American Heart Association registry found a lower proportion (55.3%), suggesting previous studies may have underestimated the diversity of cerebrovascular complications.

Haemorrhagic stroke patterns showed important temporal variations. Our meta-analysis found a 2% rate (95% CI: 0.00,0.03), but Mishra et al.’s (42) noted ICH comprised up to 40% of strokes during NYC’s pandemic peak. Shakil et al.’s (14) detailed haemorrhage subtypes (ICH 15.2%, SAH 11.4%), with Mishra et al. (42) finding 82% of ICH patients had at least one risk factor, including anticoagulation-related complications. The predominance of deep or cerebellar haemorrhages suggests mechanisms distinct from typical patterns.

Yaghi et al.’s (11) identified a high prevalence of cryptogenic strokes (65.6%) compared to contemporary (30.4%) and historical controls (25.0%). Beyrouti et al.’s (3) observation of multiple vascular territory involvement and positive antiphospholipid antibodies suggests novel COVID-19-specific mechanisms. Ntaios et al. (43) documented frequent large vessel occlusions across healthcare systems, while Grewal et al.’s (22) found 84.6% cortical involvement and high ESUS prevalence, particularly in minority populations (76.8% Latino and African-American patients). However, Shakil et al.’s (14) adjusted analysis showed no race-based differences in stroke odds.

Temporal patterns revealed crucial insights: Lodigiani et al.’s (12) found 50% of thromboembolic events occurred within 24 hours of admission, while Klok et al.’s (2) observed persistent thrombotic risk despite prophylaxis. Shakil et al. (14) reported a median of 11.5 days from symptom onset. The low incidence of haemorrhagic transformation, despite elevated D-dimer levels and widespread anticoagulation, suggests complex prothrombotic and fibrinolytic interactions.

Cavalcanti et al.’s (44) findings on cerebral venous thrombosis showed 100% mortality in patients under 41 years, with onset typically 7 days after COVID-19 symptoms, contrasting with Sluis et al.’s (20) two-week window for ischemic strokes.

These findings demonstrate that while ischemic events predominate, stroke subtypes in COVID-19 reflect complex pathophysiological mechanisms varying by demographics and timing.

### Influence of Geographical Location (Continent), Pandemic Phase (Year group), and Study Type (Multi-center, Registry-based / single-center)

Our analysis revealed relatively consistent stroke occurrence rates across geographical regions, with notable variations. Forest plot analysis demonstrated heterogeneity across continents, with North America showing the highest heterogeneity. This aligns with Shahjouei et al.’s (18) multinational study of 17,799 patients, which identified variations in stroke risk between regions, particularly noting higher rates in countries with greater healthcare expenditure. Asian studies demonstrated the lowest heterogeneity, though this might be influenced by smaller sample sizes.

Analysis across pandemic years revealed distinct patterns in reported stroke rates. Year-group analysis showed varying heterogeneity levels: substantial in 2020 (I² = 72%), increased in 2021 (I² = 90.4%), and notably decreased in 2022 (I² = 0%). This temporal variation likely reflects the evolution of COVID-19 management protocols and reporting practices. Katz et al. (10), who studied 86 COVID-19 positive stroke cases across 11 hospitals, documented that early pandemic studies were conducted under unprecedented circumstances with varying protocols. The high heterogeneity in 2021 studies potentially reflects the emergence of new viral variants and varying vaccination rates, while the lower heterogeneity in 2022 suggests a possible stabilisation in stroke reporting and management approaches.

Study methodology significantly influenced reported outcomes, as shown by subgroup analysis of study types. Multi-centre studies demonstrated the highest heterogeneity (I² = 96.8%), followed by registry-based studies (I² = 87.2%), and single-centre studies (I² = 79.1%). This variation aligns with findings from the Global COVID-19 Stroke Registry (Ntaios et al.(43)), revealing that differences in stroke ascertainment and reporting practices contributed to apparent variations in stroke rates. The higher heterogeneity in multi-centre studies might reflect greater variability in protocols and reporting standards across different institutions, as supported by Qureshi et al.’s (15) analysis of 27,676 patients across 54 healthcare facilities.

### Implications

Clinicians require robust monitoring protocols. Classification systems need standardisation. Interventions must target vulnerable groups. Key questions persist regarding stroke subtypes across variants, vaccination status, and healthcare systems. Future research should examine molecular mechanisms while standardising stroke detection methods. Population impacts and surge risks vary significantly, highlighting the need for consistent approaches.

### Strengths and Limitations

This meta-analysis presents several methodological strengths while acknowledging important limitations that warrant careful interpretation of its findings. The unprecedented scale of analysis, encompassing 97,743 patients across 17 studies for the primary forest plot provides robust statistical power and ecological validity through its multi-continental scope. However, the substantial statistical heterogeneity (I² ranging 78.7-90.4%) reflects both methodological variations and the evolving nature of the pandemic, potentially confounding temporal comparisons. While our integration of registry-based and clinical studies enabled comprehensive examination of stroke subtypes and mechanisms, the varying definitions and reporting standards across institutions may have introduced classification bias.

Furthermore, in our comparative analysis of stroke risk in COVID-19 versus non-COVID-19 hospitalised patients, the inclusion of the study by Lee et al. (27), utilised a composite outcome of major adverse cardiovascular events (MACE) and requires careful interpretation. While we extracted stroke-specific incidence rates from the Lee study’s figures, these were based on adjusted data. The comparative analysis was limited as articles compared to a non-COVID-19 group did not provide the denominator for this group, so odds ratios could not be calculated. The temporal scope spanning 2020-2022 allowed unique insights into the evolution of stroke patterns across different viral variants and treatment protocols, though this same temporal breadth complicated direct comparisons due to advancing clinical knowledge and changing healthcare behaviours. A particular strength lies in our novel analysis of age-gender interactions, revealing previously unrecognised demographic patterns in COVID-19-related stroke, yet the limited data on long-term outcomes and stroke recurrence risk constrains prognostic insights. The inclusion of both mechanically ventilated and non-ventilated patients provided crucial stratification of severity-dependent risk, though potential selection bias from early pandemic studies focusing on severe cases may have skewed initial risk estimates.

Notably, while our analysis of geographical variations offers unprecedented insight into healthcare system influences, the inability to fully account for vaccination status and viral variant impacts remains a significant limitation requiring future investigation.

## Conclusion

Our comprehensive analysis definitively establishes that COVID-19’s relationship with stroke represents a complex interplay of traditional cardiovascular risk factors and novel pathophysiological mechanisms, fundamentally challenging initial pandemic assumptions. The refined pooled stroke rate of 1%, predominantly comprising ischemic events (81%) with distinctive characteristics including high rates of cryptogenic mechanism and large vessel occlusion, suggests unique COVID-19-specific pathways requiring targeted intervention strategies. This work not only advances our understanding of COVID-19-related cerebrovascular complications but also provides an evidence-based framework for optimising stroke prevention and management protocols in future viral pandemics. The identified patterns of demographic variation, temporal evolution, and healthcare system influences demand a paradigm shift in stroke risk assessment, emphasising the need for dynamic, personalised approaches that consider both traditional risk factors and emerging viral threats. Future investigations must prioritise standardised protocols for long-term outcome assessment while exploring variant-specific pathophysiology across diverse populations, ultimately working toward more refined, evidence-based strategies for preventing and managing stroke in the context of emerging infectious diseases.

## Data Availability

All data produced in the present work are contained in the manuscript.

## Acknowledgment

The author would like to acknowledge the valuable assistance of Joanne Mcphie (Academic Liaison Librarian - Open Research & Rights) for her expert guidance in developing and refining the search terms used in this systematic review.

## References

1. Feigin VL, Stark BA, Johnson CO, Roth GA, Bisignano C, Abady GG, et al. Global, regional, and national burden of stroke and its risk factors, 1990–2019: a systematic analysis for the Global Burden of Disease Study 2019. The Lancet Neurology. 2021 Oct;20(10):795–820.

2. Klok FA, Kruip MJHA, Van Der Meer NJM, Arbous MS, Gommers DAMPJ, Kant KM, et al. Incidence of thrombotic complications in critically ill ICU patients with COVID-19. Thrombosis Research. 2020 July;191:145–7.

3. Beyrouti R, Adams ME, Benjamin L, Cohen H, Farmer SF, Goh YY, et al. Characteristics of ischaemic stroke associated with COVID-19. J Neurol Neurosurg Psychiatry. 2020 Aug;91(8):889–91.

4. Mao L, Jin H, Wang M, Hu Y, Chen S, He Q, et al. Neurologic Manifestations of Hospitalized Patients With Coronavirus Disease 2019 in Wuhan, China. JAMA Neurol. 2020 June 1;77(6):683.

5. Merkler AE, Parikh NS, Mir S, Gupta A, Kamel H, Lin E, et al. Risk of Ischemic Stroke in Patients With Coronavirus Disease 2019 (COVID-19) vs Patients With Influenza. JAMA Neurol. 2020 Nov 1;77(11):1366.

6. Nannoni S, De Groot R, Bell S, Markus HS. Stroke in COVID-19: A systematic review and meta-analysis. International Journal of Stroke. 2021 Feb;16(2):137–49.

7. Mortensen JK, Blauenfeldt RA, Hedegaard JN, Morberg Wejse C, Johnsen SP, Andersen G, et al. Prevalence and impact of SARS-CoV-2 infection among patients with acute ischaemic stroke: a nationwide register-based cohort study in Denmark. BMJ Open. 2024 Mar;14(3):e081527.

8. Nogueira RG, Abdalkader M, Qureshi MM, Frankel MR, Mansour OY, Yamagami H, et al. Global impact of COVID-19 on stroke care. International Journal of Stroke. 2021 July;16(5):573–84.

9. Srivastava PK, Zhang S, Xian Y, Xu H, Rutan C, Alger HM, et al. Acute Ischemic Stroke in Patients With COVID-19: An Analysis From Get With The Guidelines–Stroke. Stroke. 2021 May;52(5):1826–9.

10. Katz JM, Libman RB, Wang JJ, Sanelli P, Filippi CG, Gribko M, et al. Cerebrovascular Complications of COVID-19. Stroke [Internet]. 2020 Sept [cited 2025 Nov 18];51(9). Available from: https://www.ahajournals.org/doi/10.1161/STROKEAHA.120.031265

11. Yaghi S, Ishida K, Torres J, Mac Grory B, Raz E, Humbert K, et al. SARS-CoV-2 and Stroke in a New York Healthcare System. Stroke. 2020 July;51(7):2002–11.

12. Lodigiani C, Iapichino G, Carenzo L, Cecconi M, Ferrazzi P, Sebastian T, et al. Venous and arterial thromboembolic complications in COVID-19 patients admitted to an academic hospital in Milan, Italy. Thrombosis Research. 2020 July;191:9–14.

13. Tan YK, Goh C, Leow AST, Tambyah PA, Ang A, Yap ES, et al. COVID-19 and ischemic stroke: a systematic review and meta-summary of the literature. J Thromb Thrombolysis. 2020 Oct;50(3):587–95.

14. Shakil SS, Emmons-Bell S, Rutan C, Walchok J, Navi B, Sharma R, et al. Stroke Among Patients Hospitalized With COVID-19: Results From the American Heart Association COVID-19 Cardiovascular Disease Registry. Stroke. 2022 Mar;53(3):800–7.

15. Qureshi AI, Baskett WI, Huang W, Shyu D, Myers D, Raju M, et al. Acute Ischemic Stroke and COVID-19: An Analysis of 27 676 Patients. Stroke. 2021 Mar;52(3):905–12.

16. Siegler JE, Cardona P, Arenillas JF, Talavera B, Guillen AN, Chavarría-Miranda A, et al. Cerebrovascular events and outcomes in hospitalized patients with COVID-19: The SVIN COVID-19 Multinational Registry. International Journal of Stroke. 2021 June;16(4):437–47.

17. Requena M, Olivé-Gadea M, Muchada M, García-Tornel Á, Deck M, Juega J, et al. COVID-19 and Stroke: Incidence and Etiological Description in a High-Volume Center. Journal of Stroke and Cerebrovascular Diseases. 2020 Nov;29(11):105225.

18. Shahjouei S, Naderi S, Li J, Khan A, Chaudhary D, Farahmand G, et al. Risk of stroke in hospitalized SARS-CoV-2 infected patients: A multinational study. eBioMedicine. 2020 Sept;59:102939.

19. Janes F, Sozio E, Gigli GL, Ripoli A, Sbrana F, Kuris F, et al. Ischemic strokes in COVID-19: risk factors, obesity paradox, and distinction between trigger and causal association. Front Neurol. 2023 Aug 1;14:1222009.

20. Sluis WM, Linschoten M, Buijs JE, Biesbroek JM, Den Hertog HM, Ribbers T, et al. Risk, Clinical Course, and Outcome of Ischemic Stroke in Patients Hospitalized With COVID-19: A Multicenter Cohort Study. Stroke. 2021 Dec;52(12):3978–86.

21. Li Y, Li M, Wang M, Zhou Y, Chang J, Xian Y, et al. Acute cerebrovascular disease following COVID-19: a single center, retrospective, observational study. Stroke Vasc Neurol. 2020 Sept;5(3):279–84.

22. Grewal P, Pinna P, Hall JP, Dafer RM, Tavarez T, Pellack DR, et al. Acute Ischemic Stroke and COVID-19: Experience From a Comprehensive Stroke Center in Midwest US. Front Neurol. 2020 Aug 20;11:910.

23. Cantador E, Núñez A, Sobrino P, Espejo V, Fabia L, Vela L, et al. Incidence and consequences of systemic arterial thrombotic events in COVID-19 patients. J Thromb Thrombolysis. 2020 Oct;50(3):543–7.

24. Khandelwal P, Mufti FA, Tiwari AT, Singla A, Dmytriw A, Piano M, et al. Incidence, Characteristics and Outcomes of Large Vessel Stroke in COVID-19 Cohort: A Multicentric International Study. SSRN Journal [Internet]. 2020 [cited 2025 Dec 1]; Available from: https://www.ssrn.com/abstract=3617195

25. Chou SHY, Beghi E, Helbok R, Moro E, Sampson J, Altamirano V, et al. Global Incidence of Neurological Manifestations Among Patients Hospitalized With COVID-19—A Report for the GCS-NeuroCOVID Consortium and the ENERGY Consortium. JAMA Netw Open. 2021 May 11;4(5):e2112131.

26. Ramos AD, Koyfman F, Byrns K, Wu A, Yasen J, Elreda L, et al. Characterization of Hemorrhagic and Ischemic Stroke in a Diverse Cohort of COVID-19 Patients. The Neurohospitalist. 2021 Oct;11(4):295–302.

27. Lee MT, Baek MS, Kim TW, Jung SY, Kim WY. Cardiovascular outcomes between COVID-19 and non-COVID-19 pneumonia: a nationwide cohort study. BMC Med. 2023 Oct 20;21(1):394.

28. Shahjouei S, Naderi S, Li J, Chaudhary D, Griessenauer C, Farahmand G, et al. Abstract P88: Risk of Stroke in Hospitalized SARS-Cov-2 Infected Patients a Multinational Population-Based Study. Stroke [Internet]. 2021 Mar [cited 2025 Nov 18];52(Suppl_1). Available from: https://www.ahajournals.org/doi/10.1161/str.52.suppl_1.P88

29. Li Y, Li M, Wang M, Zhou Y, Chang J, Xian Y, et al. Acute cerebrovascular disease following COVID-19: a single center, retrospective, observational study. Stroke Vasc Neurol. 2020 Sept;5(3):279–84.

30. Simonsen CZ, Blauenfeldt RA, Hedegaard JN, Kruuse C, Gaist D, Wienecke T, et al. COVID-19 did not result in increased hospitalization for stroke and transient ischemic attack: A nationwide study. Euro J of Neurology. 2022 Aug;29(8):2269–74.

31. Uchino K, Kolikonda MK, Brown D, Kovi S, Collins D, Khawaja Z, et al. Decline in Stroke Presentations During COVID-19 Surge. Stroke. 2020 Aug;51(8):2544–7.

32. Nagraj S, Varrias D, Hernandez Romero G, Santos HT, Karamanis D, Sagris D, et al. Incidence of Stroke in Randomized Trials of COVID-19 Therapeutics: A Systematic Review and Meta-Analysis. Stroke. 2022 Nov;53(11):3410–8.

33. Lo Re V, Dutcher SK, Connolly JG, Perez-Vilar S, Carbonari DM, DeFor TA, et al. Association of COVID-19 vs Influenza With Risk of Arterial and Venous Thrombotic Events Among Hospitalized Patients. JAMA. 2022 Aug 16;328(7):637.

34. Akhtar N, Abid FB, Kamran S, Singh R, Imam Y, AlJerdi S, et al. Characteristics and Comparison of 32 COVID-19 and Non-COVID-19 Ischemic Strokes and Historical Stroke Patients. Journal of Stroke and Cerebrovascular Diseases. 2021 Jan;30(1):105435.

35. Dengler J, Prass K, Palm F, Hohenstein S, Pellisier V, Stoffel M, et al. Changes in nationwide in-hospital stroke care during the first four waves of COVID-19 in Germany. European Stroke Journal. 2022 June;7(2):166–74.

36. Fan S, Xiao M, Han F, Xia P, Bai X, Chen H, et al. Neurological Manifestations in Critically Ill Patients With COVID-19: A Retrospective Study. Front Neurol. 2020 July 10;11:806.

37. Topcuoglu MA, Pektezel MY, Oge DD, Bulut Yüksel ND, Ayvacioglu C, Demirel E, et al. Stroke Mechanism in COVID-19 Infection: A Prospective Case-Control Study. Journal of Stroke and Cerebrovascular Diseases. 2021 Aug;30(8):105919.

38. Qin C, Zhou L, Hu Z, Yang S, Zhang S, Chen M, et al. Clinical Characteristics and Outcomes of COVID-19 Patients With a History of Stroke in Wuhan, China. Stroke. 2020 July;51(7):2219–23.

39. Jamora RDG, Prado MB, Anlacan VMM, Sy MCC, Espiritu AI. Incidence and risk factors for stroke in patients with COVID-19 in the Philippines: An analysis of 10,881 cases. Journal of Stroke and Cerebrovascular Diseases. 2022 Nov;31(11):106776.

40. Kopp K, Motloch LJ, Lichtenauer M, Boxhammer E, Hoppe UC, Berezin AE, et al. Sex Differences in Long-Term Cardiovascular Outcomes and Mortality After COVID-19 Hospitalization During Alpha, Delta and Omicron Waves. JCM. 2024 Nov 5;13(22):6636.

41. Wilcox T, Smilowitz NR, Seda B, Xia Y, Hochman J, Berger JS. Sex Differences in Thrombosis and Mortality in Patients Hospitalized for COVID-19. The American Journal of Cardiology. 2022 May;170:112–7.

42. Mishra S, Choueka M, Wang Q, Hu C, Visone S, Silver M, et al. Intracranial Hemorrhage in COVID-19 Patients. Journal of Stroke and Cerebrovascular Diseases. 2021 Apr;30(4):105603.

43. Ntaios G, Michel P, Georgiopoulos G, Guo Y, Li W, Xiong J, et al. Characteristics and Outcomes in Patients With COVID-19 and Acute Ischemic Stroke: The Global COVID-19 Stroke Registry. Stroke [Internet]. 2020 Sept [cited 2025 Nov 18];51(9). Available from: https://www.ahajournals.org/doi/10.1161/STROKEAHA.120.031208

44. Cavalcanti DD, Raz E, Shapiro M, Dehkharghani S, Yaghi S, Lillemoe K, et al. Cerebral Venous Thrombosis Associated with COVID-19. AJNR Am J Neuroradiol. 2020 Aug;41(8):1370–6.

