## Supplementary Material for "The Effect of COVID-19 Hospitalisation on the Occurrence of Stroke: A Systematic Review and Meta-Analysis"

Supplementary Materials:

**Supplementary Table S1: Database Search Strings for Systematic Literature Review**

| **Search Database Name** | **Search string** |
|---|---|
| Web of Science Search String | TS=("COVID-19" OR "COVID-2019" OR "SARS-CoV-2" OR "SARS CoV-2" OR "Coronavirus" OR "Severe Acute Respiratory Syndrome Coronavirus-2" OR "2019-nCoV" OR "COVID-19 infection" OR "SARS- CoV-2 infection" OR "Coronavirus infection" OR "laboratory confirmed COVID-19" OR "confirmed COVID-19" OR "confirmed SARS-CoV-2" OR "COVID-19 positive" OR "SARS-CoV-2 positive" OR "confirmed 2019-nCoV" OR "confirmed Coronavirus" OR "COVID-2019 infection") AND TS=("Stroke" OR "Cerebrovascular accident" OR "CVA" OR "Brain infarction" OR "Cerebral infarction" OR "Thalamic Infarction" OR "Cerebral infarct" OR "Brain Vascular Accident" OR "Brain Vascular Accidents" OR "Apoplexy" OR "Cerebrovascular disorder" OR "Cerebrovascular event" OR "Brain ischemia" OR "Cerebral ischemia" OR "Intracranial haemorrhage" OR "Cerebrovascular disease" OR "Cerebrovascular insult" OR "Brain attack" OR "Cerebral thrombosis" OR "Cerebral embolism" OR "Subarachnoid hemorrhage" OR "Intracerebral hemorrhage") AND TS=("hospitalisation" OR "hospitalization" OR "inpatient" OR "admission") |
| PubMed Search String | (("COVID-19"[Title/Abstract] OR "COVID-2019"[Title/Abstract] OR "SARS-CoV-2"[Title/Abstract] OR "SARS CoV-2"[Title/Abstract] OR "Coronavirus"[Title/Abstract] OR "Severe Acute Respiratory Syndrome Coronavirus-2"[Title/Abstract] OR "2019-nCoV"[Title/Abstract] OR "COVID-19 infection"[Title/Abstract] OR "SARS-CoV-2 infection"[Title/Abstract] OR "Coronavirus<br><br>infection"[Title/Abstract] OR "laboratory confirmed COVID-19"[Title/Abstract] OR "confirmed COVID-19"[Title/Abstract] OR "confirmed SARS-CoV-2"[Title/Abstract] OR "COVID-19 positive"[Title/Abstract] OR "SARS-CoV-2 positive"[Title/Abstract] OR "confirmed 2019- nCoV"[Title/Abstract] OR "confirmed Coronavirus"[Title/Abstract] OR "COVID-2019 infection"[Title/Abstract])) AND ("Stroke"[Title/Abstract] OR "Cerebrovascular<br><br>accident"[Title/Abstract] OR "CVA"[Title/Abstract] OR "Brain infarction"[Title/Abstract] OR "Cerebral infarction"[Title/Abstract] OR "Thalamic Infarction"[Title/Abstract] OR "Cerebral infarct"[Title/Abstract] OR "Brain Vascular Accident"[Title/Abstract] OR "Brain Vascular<br><br>Accidents"[Title/Abstract] OR "Apoplexy"[Title/Abstract] OR "Cerebrovascular disorder"[Title/Abstract] OR "Cerebrovascular |

| | |
|---|---|
| | event"[Title/Abstract] OR "Brain ischemia"[Title/Abstract] OR "Cerebral ischemia"[Title/Abstract] OR "Intracranial haemorrhage"[Title/Abstract] OR "Cerebrovascular disease"[Title/Abstract] OR<br><br>"Cerebrovascular insult"[Title/Abstract] OR "Brain attack"[Title/Abstract] OR "Cerebral thrombosis"[Title/Abstract] OR "Cerebral embolism"[Title/Abstract] OR "Subarachnoid hemorrhage"[Title/Abstract] OR "Intracerebral hemorrhage"[Title/Abstract]) AND<br><br>("hospitalisation"[Title/Abstract] OR "hospitalization"[Title/Abstract] OR "inpatient"[Title/Abstract] OR "admission"[Title/Abstract]) ) |
| Scopus Search String | TITLE-ABS-KEY(("COVID-19" OR "COVID-2019" OR "SARS-CoV-2" OR "SARS CoV-2" OR<br><br>"Coronavirus" OR "Severe Acute Respiratory Syndrome Coronavirus-2" OR "2019-nCoV" OR "COVID-19 infection" OR "SARS-CoV-2 infection" OR "Coronavirus infection" OR "laboratory confirmed COVID-19" OR "confirmed COVID-19" OR "confirmed SARS-CoV-2" OR "COVID-19 positive" OR "SARS-CoV-2 positive" OR "confirmed 2019-nCoV" OR "confirmed Coronavirus" OR "COVID-2019 infection")) AND TITLE-ABS-KEY(("Stroke" OR "Cerebrovascular accident" OR "CVA" OR "Brain infarction" OR "Cerebral infarction" OR "Thalamic Infarction" OR "Cerebral infarct" OR "Brain Vascular Accident" OR "Brain Vascular Accidents" OR "Apoplexy" OR "Cerebrovascular disorder" OR "Cerebrovascular event" OR "Brain ischemia" OR "Cerebral<br><br>ischemia" OR "Intracranial haemorrhage" OR "Cerebrovascular disease" OR "Cerebrovascular insult" OR "Brain attack" OR "Cerebral thrombosis" OR "Cerebral embolism" OR "Subarachnoid hemorrhage" OR "Intracerebral hemorrhage")) AND TITLE-ABS-KEY(("hospitalisation" OR<br><br>"hospitalization" OR "inpatient" OR "admission")) |

Note: Search queries were executed in Web of Science, PubMed, and Scopus without language restrictions.

**Supplementary Table S2**: Quality Assessment of Included Studies Cohort Studies Using the Newcastle-Ottawa Scale

| First Author (Year) | Selection (Max 4★) | Comparability (Max 2★) | Outcome (Max 3★) | Total Score |
|---|---|---|---|---|
| Shakil SS (2021) | ★★★★ | ★★ | ★★★ | 9/9 |
| Qureshi AI (2020) | ★★★★ | ★★ | ★★ | 8/9 |
| Merkler AE (2020) | ★★★★ | ★★ | ★★ | 8/9 |
| Requena M (2020) | ★★★ | ★ | ★★ | 6/9 |
| Lee M-T (2023) | ★★★★ | ★★ | ★★ | 7/9 |
| Siegler JE (2020) | ★★★★ | ★★ | ★★ | 8/9 |
| Sluis WM (2021) | ★★★★ | ★★ | ★★ | 8/9 |
| Yaghi S (2020) | ★★★★ | ★★ | ★★ | 8/9 |
| Li Y (2020) | ★★★ | ★ | ★★ | 6/9 |
| Cantador (2020) | ★★★ | ★★ | ★★ | 7/9 |
| Lodigiani (2020) | ★★★ | ★★ | ★★ | 7/9 |
| Mao (2020) | ★★★ | ★ | ★★ | 6/9 |
| Shahjouei (2020) | ★★★★ | ★★ | ★★★ | 9/9 |
| Chou SHY (2021) | ★★★ | ★★ | ★★ | 7/9 |
| Katz JM (2020) | ★★★★ | ★★ | ★★ | 8/9 |
| Grewal P (2020) | ★★★ | ★ | ★★ | 6/9 |
| Janes F (2023) | ★★★★ | ★★ | ★★ | 8/9 |
| Khandelwal P (2021) | ★★★ | ★★ | ★ | 6/9 |
| Ramos AD (2021) | ★★★★ | ★ | ★★ | 7/9 |

Note: A study can be awarded a maximum of one star (★) for each numbered item within the Selection and Outcome categories. A maximum of two stars can be given for Comparability. **Scoring Key:** Selection domain: Maximum 4 stars (★); Comparability domain: Maximum 2 stars (★); Outcome domain: Maximum 3 stars (★). Total maximum score is 9 stars. **Quality Categories:** High quality: 8–9 stars; Moderate quality: 6–7 stars; Low quality: ≤5 stars. **Abbreviations:** NOS, Newcastle-Ottawa Scale.

**Supplementary Table S3: Newcastle-Ottawa Scale Scoring Criteria Guide for COVID-19 and Stroke Studies**

| Domain | Criterion | Scoring Options | Star Award Criteria |
|---|---|---|---|
| SELECTION | | | Maximum 4★ total (max 1★ per criterion) |
| | 1. Representativeness of exposed cohort | | Award 1★ if (a) OR (b): |
| | | a) Truly representative of hospitalised COVID-19 patients (consecutive admissions, multicenter studies, nationwide registries) | ★ |
| | | b) Somewhat representative of hospitalised COVID-19 patients (single-center studies with clear inclusion criteria) | ★ |
| | | c) Selected group of COVID-19 patients (only ICU patients, specific age groups) | 0 |
| | | d) No description of cohort derivation | 0 |
| | 2. Selection of non-exposed cohort | | Award 1★ if (a) only: |
| | | a) Drawn from same community as exposed cohort (COVID-19 patients without stroke or non-COVID stroke patients during same period) | ★ |
| | | b) Drawn from different source | 0 |
| | | c) No description of non-exposed cohort derivation | 0 |

| | | | |
|---|---|---|---|
| | 3. Ascertainment of exposure | | Award 1★ if (a) OR (b): |
| | | a) Secure record (laboratory-confirmed SARS-CoV-2 infection, hospital records) | ★ |
| | | b) Structured interview | ★ |
| | | c) Written self-report | 0 |
| | | d) No description | 0 |
| | 4. Demonstration that outcome was not present at study start | | Award 1★ if (a) only: |
| | | a) Yes (new stroke during hospitalisation, exclusion of prior stroke history) | ★ |
| | | b) No | 0 |
| COMPARABILITY | | | Maximum 2★ total |
| | 1. Comparability of cohorts based on design/analysis | | Award up to 2★: |
| | | a) Study controls for age | +1★ |
| | | b) Study controls for additional vascular risk factors (hypertension, diabetes, cardiovascular disease) | +1★ |
| OUTCOME | | | Maximum 3★ total (max 1★ per criterion) |

| | | | |
|---|---|---|---|
| | 1. Assessment of outcome | | Award 1★ if (a) OR (b): |
| | | a) Independent blind assessment | ★ |
| | | b) Record linkage (confirmed stroke diagnosis through imaging/clinical records) | ★ |
| | | c) Self-report | 0 |
| | | d) No description | 0 |
| | 2. Follow-up duration adequate for outcomes | | Award 1★ if (a) only: |
| | | a) Yes (minimum hospital stay duration or 30-day follow-up) | ★ |
| | | b) No | 0 |
| | 3. Adequacy of follow-up of cohorts | | Award 1★ if (a) OR (b): |
| | | a) Complete follow-up - all subjects accounted for | ★ |
| | | b) Subjects lost to follow-up unlikely to introduce bias (≤10% lost, or description provided) | ★ |
| | | c) Follow-up rate <90% and no description of those lost | 0 |
| | | d) No statement | 0 |

Note: Quality Assessment Categories: High Quality: 8-9 stars, Moderate Quality: 6-7 stars and Low Quality: ≤5 stars. Special Considerations for COVID-19 and Stroke Studies: Exposure ascertainment should include confirmed SARS-CoV-2 infection, Outcome assessment should include validated stroke diagnosis, Follow-up period should cover at least the hospital stay duration, Adjustment for COVID-19 severity is considered an important additional factor and Clear documentation of stroke timing relative to COVID-19 diagnosis is important.

Raw Data Extraction

**Supplementary Table S4: Characteristics and Quantitative Findings of Included Studies Investigating Stroke Occurrence in Hospitalised COVID-19 Patients.**

| Authors | Study Type | Study Period | Location | Sample Size (N) | Population | Effect Estimate | 95% CI | P value |
|---|---|---|---|---|---|---|---|---|
| Chou SHY et al. (2021) | Cohort study | Mar-Sep 2020 | 28 centers across 13 countries | 3,054 | Hospitalised COVID-19 patients | 0.03 | 0.02-0.04 | <0.001 |
| Shakil SS (2021) | Retrospective cohort | Jan-Nov 2020 | USA | 21,073 | Hospitalised COVID-19 patients | 0.01 | 0.01-0.02 | <0.05 |
| Qureshi AI | Retrospective cohort | Dec 2019-Apr 2020 | USA | 8,163 | Hospitalised patients | 0.01 | 0.01-0.02 | <0.0001 |
| Katz (2020) | Retrospective case series | Mar-Apr 2020 | USA | 10,596 | Hospitalised COVID-19 patients | 0.01 | 0.01-0.01 | <0.001 |
| Siegler et al | Retrospective cohort | Feb-Jun 2020 | Multinational | 14,483 | Hospitalised COVID-19 patients | 0.01 | 0.01-0.01 | <0.001 |
| Requena M | Retrospective cohort | Mar-Apr 2020 | Spain | 2,050 | Hospitalised COVID-19 patients | 0.01 | 0.01-0.02 | 0.04 |
| Shahjouei et al (2020) | Multicenter retrospective | Up to May 2020 | 11 countries | 17,799 | Hospitalised COVID-19 patients | 0.01 | 0.01-0.01 | <0.001 |
| Janes et al (2023) | Retrospective cohort | Feb-Dec 2020 | Philippines | 4,105 | Hospitalised COVID-19 patients | 0.02 | 0.01-0.02 | <0.0001 |
| Sluis et al (2021) | Multicenter cohort | Mar-Aug 2020 | Netherlands | 2,147 | Hospitalised COVID-19 patients | 0.02 | 0.01-0.02 | <0.001 |

| Authors | Study Type | Study Period | Location | Sampl e Size (N) | Populatio n | Effect Estima t e | 95% CI | P value |
|---|---|---|---|---|---|---|---|---|
| Li Y | Single- center retrospective | Jan-Feb 2020 | China | 219 | Hospitalised COVID-19 patients | 0.05 | 0.03-0.09 | 0.009 |
| Lodigiani | Retrospective cohort | Feb-Apr 2020 | Italy | 388 | Hospitalised COVID-19 patients | 0.02 | 0.01-0.04 | <0.001 |
| Mao et al (2020) | Retrospective observational | Jan-Feb 2020 | China | 214 | Hospitalised COVID-19 patients | 0.03 | 0.01-0.06 | 0.03 |
| Yaghi et al (2020) | Retrospective cohort | Mar-Apr 2020 | USA | 3,556 | Hospitalised COVID-19 patients | 0.01 | 0.01-0.01 | <0.0001 |
| Grewal et al (2020) | Retrospective cohort | Not provid e d | Not provided | 650 | Hospitalised COVID-19 patients | 0.02 | 0.01-0.03 | Not provided |
| Cantador | Retrospective observation al | Feb-Apr 2020 | Spain | 1,419 | Hospitalised COVID-19 patients | 0.01 | 0.00-0.01 | 0.005 |
| Merkler AE | Retrospective cohort | Mar-May 2020 | USA | 1,683 | Hospitalised patients | 0.02 | 0.01-0.03 | <0.05 |
| Ramos et al. (2021) | Retrospective observation al | Mar-May 2020 | USA | 2,401 | Hospitalised COVID-19 patients | 0.015 | Not reporte d | <0.0001 |
| Khandelw al et al (2021) | Cross-sectional retrospective | Mar-May 2020 | USA, UK, Spain, Italy | 6,698 | Hospitalised COVID-19 patients | 0.013 | 0.0075-0.017 | Not reported |
| Lee et al. (2023) | Nationwide cohort | Oct 2020- | Korea | 132,78 4 | Hospitalised COVID-19 patients | 0.01 | 0.01-0.02 | Not significan t |

| | | | | | | | | |
|---|---|---|---|---|---|---|---|---|
| | | Sep 2021 | | | | | | |
| Chou SHY et al. (2021) | Cohort study | Mar-Sep 2020 | 28 centers across 13 countries | 3,054 | Hospitalised COVID-19 patients | 0.03 | 0.02-0.04 | <0.001 |

Abbreviations: CI, confidence interval; N, total sample size.

Note**:** Effect estimates represent the proportion (prevalence) of stroke occurrence reported in each study. P-values indicate the statistical significance of the reported occurrence where available.

**Supplementary Table S5: Calculated Odds Ratio's and Raw data for the COVID-19 Hospitalised vs non-COVID-19 Control group Forest Plot**

| Study ID | COVID-19 Stroke Cases | COVID-19 No Stroke | Control Stroke Cases | Control No Stroke | Odds Ratio | 95% CI |
|---|---|---|---|---|---|---|
| Merkler 2020 | 31 | 1,885 | 3 | 1,483 | 7.6 | 2.3-25.2 |
| Lee 2023 | 112 | 125,698 | 42 | 28,450 | 0.84 | 0.69-1.03 |
| Ramos 2021 | 37 | 2,364 | 47 | 1,468 | 0.49 | Not reported |

Abbreviations: CI, confidence interval; OR, odds ratio.

Note: The table presents the raw event data and calculated odds ratios used to compare stroke occurrence between hospitalised COVID-19 patients and non-COVID-19 control groups. "Not reported" indicates that the confidence interval was not explicitly provided in the primary source data for that specific metric.

**Supplementary Table S6. Quantitative Data for the Subgroup Analysis Examining the Association Between Mechanical Ventilation and Stroke Risk in Hospitalised COVID-19 Patients.**

| Study ID | Stroke with MV (A) | Stroke without MV (B) | No Stroke with MV (C) | No Stroke without MV (D) | Odds Ratio | log(OR) | SE | 95% CI |
|---|---|---|---|---|---|---|---|---|
| Shahjouei et al. | 31 | 53 | 428 | 2,628 | 3.59 | 1.28 | 0.232 | 2.27-5.67 |
| Merkler et al. | 11 | 20 | 319 | 1,566 | 2.70 | 0.993 | 0.352 | 1.35-5.39 |
| Qureshi et al. | 7 | 96 | 265 | 7,341 | 2.02 | 0.703 | 0.403 | 0.92-4.45 |
| Shakil et al. | 93 | 67 | 3,931 | 16,853 | 5.95 | 1.783 | 0.137 | 4.55-7.78 |

Abbreviations: MV, mechanical ventilation; OR, odds ratio; SE, standard error.

Note: The table presents the raw event data, calculated odds ratios, standard errors, and statistical weights (fixed and random effects) used for the meta-analysis comparing stroke risk between patients requiring mechanical ventilation and those who did not.

**Supplementary Table S7. Quantitative Data for the Subgroup Analysis Examining the Association Between Gender (Male vs. Female) and Stroke Risk in Hospitalised COVID-19 Patients.**

| Study | Total COVID-19 Patients | Males with Stroke | Females with Stroke | Males without Stroke | Females without Stroke | Calculated OR (95% CI) |
|---|---|---|---|---|---|---|
| Sluis et al (2021) | 2,147 | 22 | 16 | 1,356 | 753 | 0.76 (0.40-1.46) |
| Janes et al (2023) | 219 | 6 | 5 | 83 | 125 | 0.81 (0.44-1.47) |
| Qureshi et al (2021) | 8,163 | 46 | 57 | 3,575 | 4,031 | 0.91 (0.62-1.34) |
| Shakil et al (2021) | 21,073 | 101 | 59 | 11,208 | 9,576 | 1.46 (1.06-2.02) |
| Shahjouei et al (2020) | 17,799 | 109 | 47 | 10,214 | 7,429 | 1.69 (1.20-2.38) |
| Li et al (2020) | 219 | 6 | 5 | 83 | 125 | 1.81 (0.53-6.12) |

**Abbreviations:** CI, confidence interval; LogOR, logarithm of the odds ratio; SE, standard error. **Note:** The table presents the raw event data stratified by gender, alongside calculated log-odds ratios, standard errors, and statistical weights (fixed and random effects) used for the meta-analysis comparing stroke risk between male and female patients.

**Supplementary Table S8. Quantitative Data for the Subgroup Analysis Examining the Association Between Intensive Care Unit (ICU) Admission and Stroke Risk in Hospitalised COVID-19 Patients.**

| Study ID | Stroke with ICU (A) | Stroke without ICU (B) | No Stroke with ICU (C) | No Stroke without ICU (D) | Odds Ratio [95% CI] |
|---|---|---|---|---|---|
| Shahjouei et al (2020) | 110 | 50 | 6,270 | 19,745 | 6.93 [4.95-9.69] |
| Lodigiani et al (2020) | 3 | 6 | 58 | 321 | 2.77 [0.67-11.38] |
| Merkler et al (2020) | 19 | 12 | 455 | 1,430 | 4.98 [2.40-10.33] |
| Shakil et al (2021) | 215 | 74 | 6,270 | 14,514 | 6.73 [5.16-8.77] |

Abbreviations: ICU, intensive care unit; LogOR, logarithm of the odds ratio; SE, standard error.

Note: The table presents the raw event data stratified by ICU admission status, alongside calculated log-odds ratios, standard errors, and statistical weights (fixed and random effects) used for the meta-analysis comparing stroke risk between patients admitted to the ICU and those who were not.

Comorbidities:

**Supplementary Table S9: Quantitative Data for the Subgroup Analysis Examining the Association Between Hypertension (HTN) and Stroke Risk in Hospitalised COVID-19 Patients.**

| Study ID | Stroke with HTN | Stroke without HTN | No Stroke with HTN | No Stroke without HTN | Calculated OR | 95% CI | Log(O R) | SE |
|---|---|---|---|---|---|---|---|---|
| Shahjoue i 2020 | 61 | 33 | 1912 | 2640 | 2.55 | 1.65-3.94 | 0.936 | 0.222 |
| Qureshi 2021 | 87 | 16 | 3662 | 3944 | 5.87 | 3.42-10.07 | 1.77 | 0.275 |
| Shakil 2022 | 129 | 31 | 12139 | 8645 | 2.96 | 2.00-4.39 | 1.085 | 0.200 |
| Sluis 2021 | 14 | 24 | 1020 | 1089 | 0.62 | 0.32-1.21 | -0.478 | 0.340 |

Abbreviations: HTN, hypertension; LogOR, logarithm of the odds ratio; SE, standard error. Note: The table presents the raw event data stratified by hypertension status, alongside calculated log-odds ratios, standard errors, and statistical weights (fixed and random effects) used for the meta-analysis comparing stroke risk between patients with and without hypertension.

**Supplementary Table S10. Quantitative Data for the Subgroup Analysis Examining the Association Between Diabetes Mellitus and Stroke Risk in Hospitalised COVID-19 Patients.**

| Study ID | Stroke with DM | Stroke without DM | No Stroke with DM | No Stroke without DM | Calculated OR | 95 % CI | Log(OR) | SE |
|---|---|---|---|---|---|---|---|---|
| Shahjouei 2020 | 32 | 62 | 1312 | 3376 | 1.33 | 0.86 - 2.05 | 0.285 | 0.221 |
| Qureshi 2021 | 58 | 45 | 2295 | 5311 | 2.98 | 2.01 - 4.42 | 1.09 | 0.201 |
| Shakil 2022 | 72 | 88 | 7300 | 13484 | 1.51 | 1.11 - 2.06 | 0.412 | 0.159 |
| Sluis 2021 | 8 | 30 | 560 | 1549 | 0.74 | 0.34 - 1.61 | -0.301 | 0.397 |

Abbreviations: DM, diabetes mellitus; LogOR, logarithm of the odds ratio; SE, standard error. Note: The table presents the raw event data stratified by diabetes status, alongside calculated log-odds ratios, standard errors, and statistical weights (fixed and random effects) used for the meta-analysis comparing stroke risk between patients with and without diabetes mellitus.

**Supplementary Table S11. Quantitative Data for the Subgroup Analysis Examining the Association Between Ischemic Heart Disease or Coronary Artery Disease and Stroke Risk in Hospitalised COVID-19 Patients.**

| Study ID | Stroke with IHD/CAD | Stroke without IHD/CAD | No Stroke with IHD/CAD | No Stroke without IHD/CAD | Calculated OR | 95% CI | Log(OR) | SE |
|---|---|---|---|---|---|---|---|---|
| Shahjou ei 2020 | 28 | 66 | 560 | 4112 | 3.11 | 1.97 - 4.91 | 1.135 | 0.234 |
| Qureshi 2021 | 24 | 79 | 2026 | 5580 | 0.84 | 0.53 - 1.32 | -0.174 | 0.232 |
| Shakil 2022 | 24 | 136 | 2026 | 18758 | 1.63 | 1.05 - 2.53 | 0.489 | 0.224 |
| Sluis 2021 | 5 | 33 | 443 | 1666 | 0.57 | 0.22 - 1.47 | -0.562 | 0.483 |

Abbreviations: CAD, coronary artery disease; CI, confidence interval; IHD, ischemic heart disease; LogOR, logarithm of the odds ratio; OR, odds ratio; SE, standard error. Note: The table presents the raw event data stratified by the presence of ischemic heart disease/coronary artery disease, alongside calculated log-odds ratios, standard errors, and statistical weights (fixed and random effects) used for the meta-analysis comparing stroke risk between patients with and without these conditions.

**Supplementary Table S12. Quantitative Data for the Subgroup Analysis Examining the Association Between Atrial Fibrillation and Stroke Risk in Hospitalised COVID-19 Patients.**

| Study ID | Strok e with AF | Stroke without AF | No Stroke with AF | No Stroke without AF | Calculated OR | 95% CI | Log(OR) | SE |
|---|---|---|---|---|---|---|---|---|
| Shahjoue i 2020 | 9 | 85 | 178 | 2366 | 1.41 | 0.70 - 2.84 | 0.344 | 0.35 8 |
| Qureshi 2021 | 29 | 74 | 768 | 6838 | 3.48 | 2.24 - 5.41 | 1.25 | 0.22 5 |
| Shakil 2022 | 31 | 129 | 1977 | 18807 | 2.28 | 1.54 - 3.38 | 0.824 | 0.20 1 |
| Sluis 2021 | 4 | 34 | 278 | 1831 | 0.77 | 0.27 - 2.20 | -0.261 | 0.53 4 |

Abbreviations: AF, atrial fibrillation; CI, confidence interval; LogOR, logarithm of the odds ratio; OR, odds ratio; SE, standard error. Note: The table presents the raw event data stratified by the presence of atrial fibrillation, alongside calculated odds ratios, log-odds ratios, and standard errors used for the meta-analysis comparing stroke risk between patients with and without atrial fibrillation.

**Supplementary Table S13. Quantitative Data for the Prevalence of Stroke Sub-Types Among Hospitalised COVID-19 Patients with Stroke**

| Stroke Type | Study | Events |
|---|---|---|
| **Ischemic Stroke** | | |
| Shakil et al., 2022 | 160 | 254 |
| Shahjouei et al., 2020 | 123 | 156 |
| Siegler et al., 2021 | 156 | 172 |
| Mao et al., 2020 | 5 | 6 |
| Li et al., 2020 | 10 | 11 |
| Requena et al., 2020 | 21 | 25 |
| Katz et al., 2020 | 72 | 86 |
| **Hemorrhagic Stroke** | | |
| Shakil et al., 2022 | 98* | 254 |
| Shahjouei et al., 2020 | 27** | 156 |
| Siegler et al., 2021 | 28 | 172 |
| Mao et al., 2020 | 1 | 6 |
| Li et al., 2020 | 1 | 11 |
| Requena et al., 2020 | 4 | 25 |
| Katz et al., 2020 | 14 | 86 |
| **CVST** | | |
| Shakil et al., 2022 | 2 | 254 |
| Shahjouei et al., 2020 | 6 | 156 |
| Siegler et al., 2021 | 3 | 172 |

Abbreviations: CVST, cerebral venous sinus thrombosis; n, number of events for the specific sub-type; N, total number of stroke patients included in the study analysis. Note: The table presents the raw counts of specific stroke sub-types relative to the total number of stroke events recorded in each study.

Forest Plots

**Supplementary Figure S1:**

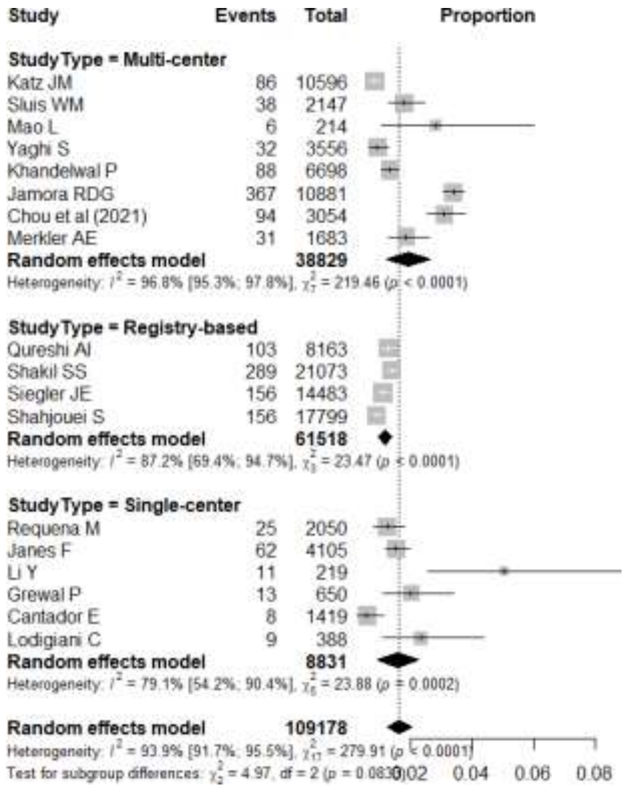

**Supplementary Figure S1: Forest Plot of Stroke Occurrence in Hospitalised COVID-19 Patients Stratified by Study Design.** This sensitivity analysis subgrouped studies into Multi-center, Registry-based, and Single-center categories to investigate sources of heterogeneity. The forest plot includes a total population of 109,178 patients. Individual study point estimates for stroke proportion are represented by grey squares, with 95% confidence intervals (CI) indicated by horizontal lines. The size of each square corresponds to the study's weight in the random-effects analysis. The diamonds represent the pooled proportion for each subgroup and the overall model. The analysis reveals substantial heterogeneity across all designs, highest in multi-center studies ($I^2$ = 96.8%), followed by registry-based ($I^2$ = 87.2%) and single-center studies ($I^2$ = 79.1%). The test for subgroup differences (p = 0.08) indicates that while study design contributes to variance, it does not fully explain the heterogeneity observed in stroke occurrence rates.

**Supplementary Figure S2:**

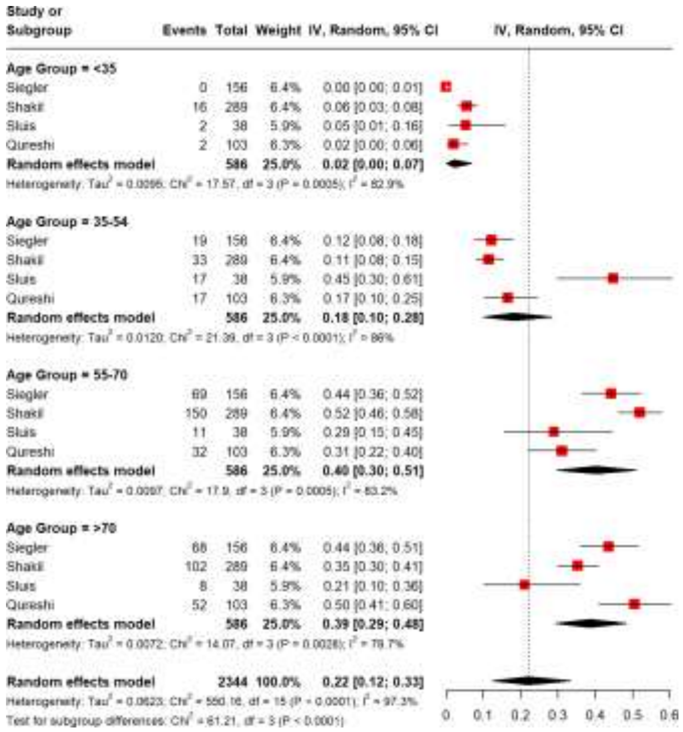

**Supplementary Figure S2: Forest Plot of Stroke Occurrence in Hospitalised COVID-19 Patients Stratified by Age Group.** This subgroup analysis examines the distribution of stroke events across four distinct age categories (<35, 35–54, 55–70, and >70 years) to identify age-dependent patterns in stroke risk. The analysis aggregates data from four studies, comprising a total population of 2,344 patients. Red squares represent the point estimates of stroke proportion for individual studies within each age band, with horizontal lines indicating the 95% confidence intervals (CI). The black diamonds represent the pooled proportion for each subgroup. The results demonstrate a clear age-dependent trend: stroke occurrence was lowest in patients <35 years (2%, 95% CI: 0.00,0.07) and rose significantly in the middle age groups, peaking in patients aged 55–70 (40%, 95% CI:0.30,0.51) and those >70 years (39%, 95% CI: 0.29,0.48). Significant heterogeneity was observed across all age groups ($I^2$ range: 78.7%-86%), and the test for subgroup differences ($X^2$= 61.21, p < 0.0001) confirmed that stroke proportions varied significantly by age.

**Supplementary Figure S3**:

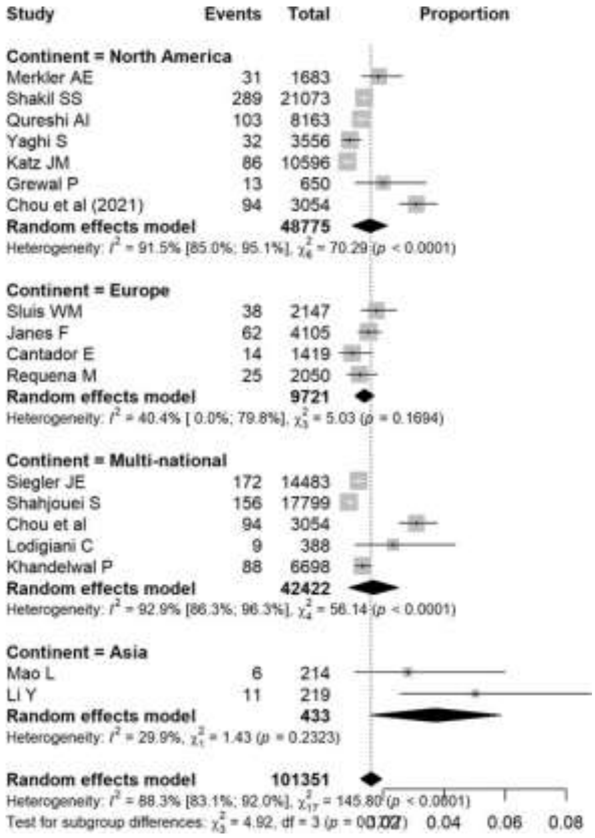

**Supplementary Figure S3: Geographical Sub-group, Forest Plot of Stroke Occurrence in Hospitalised COVID-19 Patients Stratified by Geographical Region**. This sensitivity analysis subgroups the included studies by continent (North America, Europe, Asia) and Multi-national collaborations to assess potential geographical variations in stroke risk. The analysis encompasses a total population of 101,351 patients. Grey squares represent the point estimates of stroke proportion for individual studies, with the size of each square proportional to its weight in the random-effects model. Horizontal lines indicate the 95% confidence intervals (CI). The black diamonds represent the pooled proportion for each geographical subgroup and the overall total. The results demonstrate substantial heterogeneity in North American ($I^2$ = 91.5%) and Multi-national studies ($I^2$ = 92.9%), whereas European ($I^2$ = 40.4%) and Asian ($I^2$ = 29.9%) cohorts exhibited lower heterogeneity. The test for subgroup differences ($X^2$ = 4.92, p = 0.18) indicates that stroke occurrence did not differ significantly across these geographical regions.

**Supplementary Figure S4**:

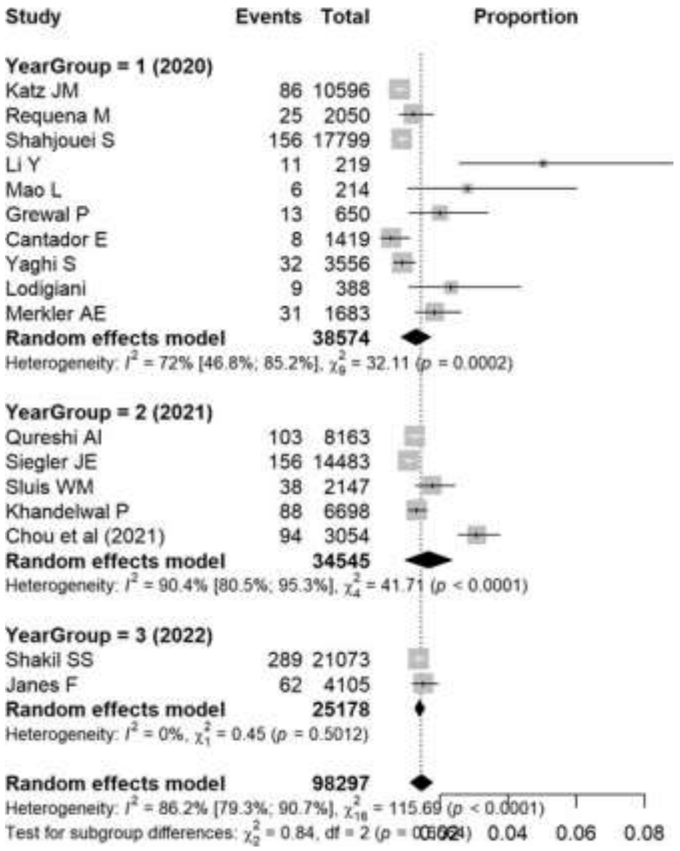

**Supplementary Figure S4: Forest Plot of Stroke Occurrence in Hospitalised COVID-19 Patients Stratified by Pandemic Phase (Year).** This sensitivity analysis subgroups the included studies by year (Year Group 1 = 2020, Year Group 2 = 2021, Year Group 3 = 2022) to investigate temporal variations in reported stroke risk and heterogeneity. The analysis comprises a total population of 98,297 patients. Grey squares represent the point estimates of stroke proportion for individual studies, with the size of each square proportional to its weight in the random-effects model. Horizontal lines indicate the 95% confidence intervals (CI). The black diamonds represent the pooled proportion for each year group and the overall total. The analysis highlights a reduction in heterogeneity over time: while 2020 and 2021 showed substantial heterogeneity ($I^2$ = 72% and $I^2$ = 90.4%, respectively), the 2022 subgroup demonstrated no heterogeneity ($I^2$ = 0%). The test for subgroup differences ($X^2$ = 0.84, df = 2, p = 0.66) indicates that the pooled stroke occurrence rate did not differ significantly across the three years.

**Supplementary Figure S5:**

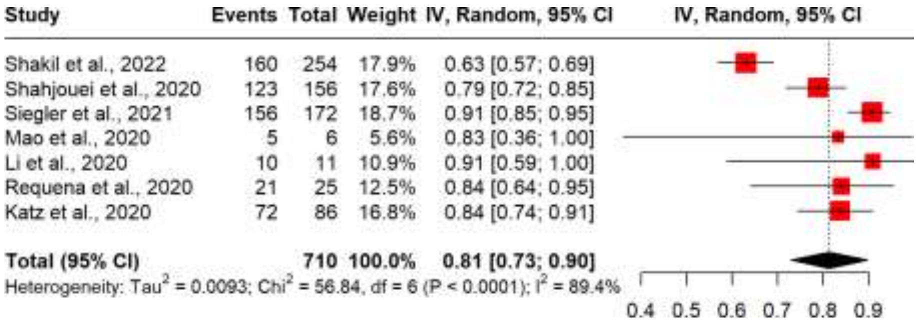

**Supplementary Figure S5: Forest Plot of Ischemic Stroke Prevalence Among Hospitalised COVID-19 Patients with Stroke.** This subgroup meta-analysis assesses the proportion of ischemic stroke cases relative to the total number of stroke events documented in hospitalised COVID-19 patients. The analysis aggregates data from 7 studies, comprising a total of 710 stroke patients. Red squares represent the point estimates for the proportion of ischemic strokes in each study, with horizontal lines indicating the 95% confidence intervals (CI). The size of each square corresponds to the study's weight in the random-effects analysis. The black diamond represents the overall pooled proportion of 81% (0.81; 95% CI: 0.73,0.90), confirming ischemic stroke as the predominant subtype. Significant statistical heterogeneity was observed across the included studies ($I^2$ = 89.4%, $p < 0.0001$).

**Supplementary Figure S6:**

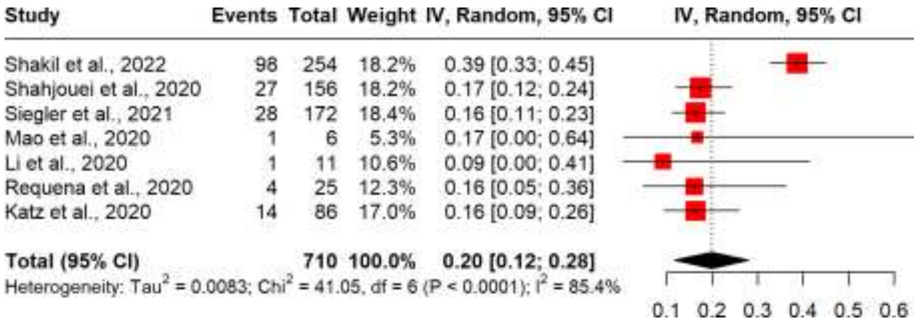

**Supplementary Figure S6: Forest Plot of Haemorrhagic Stroke Prevalence Among Hospitalised COVID-19 Patients with Stroke**. This subgroup meta-analysis examines the proportion of haemorrhagic stroke cases relative to the total number of stroke events recorded in hospitalised COVID-19 patients. The analysis synthesises data from 7 studies, encompassing a total population of 710 stroke patients. Red squares denote the point estimates for the proportion of haemorrhagic strokes within each study, with horizontal lines representing the 95% confidence intervals (CI). The size of each square is proportional to the weight assigned to the study in the random-effects model. The black diamond indicates the overall pooled proportion of 20% (0.20; 95% CI: 0.12,0.28). Significant statistical heterogeneity was detected among the studies ($I^2$ = 85.4%, $p < 0.0001$).

**Supplementary Figure S7:**

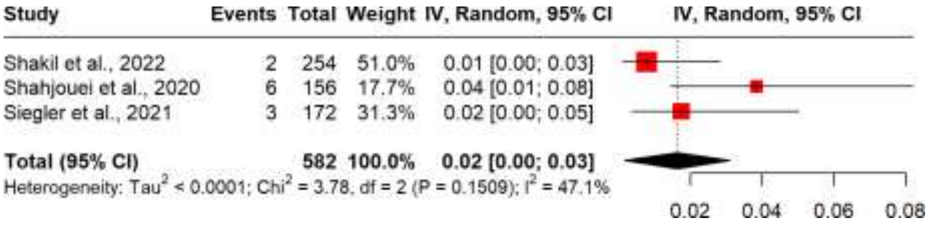

**Supplementary Figure S7: Forest Plot of Cerebral Venous Sinus Thrombosis (CVST) Prevalence Among Hospitalised COVID-19 Patients with Stroke.** This subgroup meta-analysis evaluates the proportion of Cerebral Venous Sinus Thrombosis (CVST) cases relative to the total number of stroke events documented in hospitalised COVID-19 patients. The analysis aggregates data from 3 studies, comprising a total population of 582 stroke patients. Red squares represent the point estimates for the proportion of CVST in each study, with horizontal lines indicating the 95% confidence intervals (CI). The size of each square corresponds to the study's weight in the random-effects analysis. The black diamond represents the overall pooled proportion of 2% (0.02; 95% CI: 0.00,0.03). Moderate heterogeneity was observed among the studies ($I^2$ = 47.1%, p = 0.15).

Funnel Plots

**Supplementary Figure S8:**

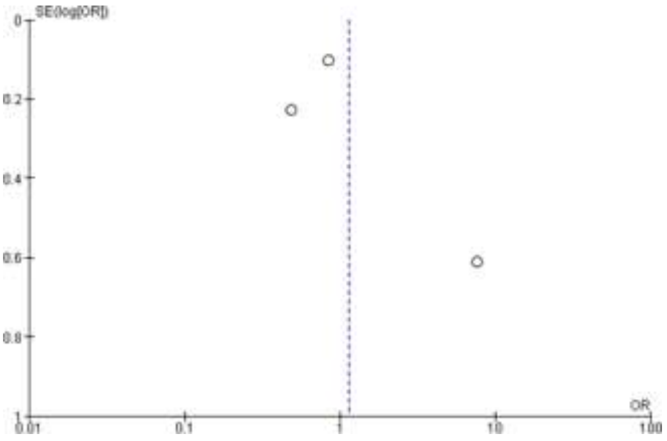

**Supplementary Figure S8:Publication Bias Assessment**. The funnel plot for the COVID-19 Hospitalisation and Stroke Risk Compared to Non-COVID-19 Hospitalised Controls comparative analysis shows asymmetric distribution of the three included studies**,** with points scattered on both sides of the null effect line (OR = 1). The limited number of studies and their asymmetric distribution, combined with high heterogeneity ($I^2$ =89%, p = 0.0001), suggests potential publication bias but makes definitive assessment challenging. The temporal pattern of effect sizes (2020-2023) may reflect evolving clinical practices rather than publication bias alone.

**Supplementary Figure S9**:

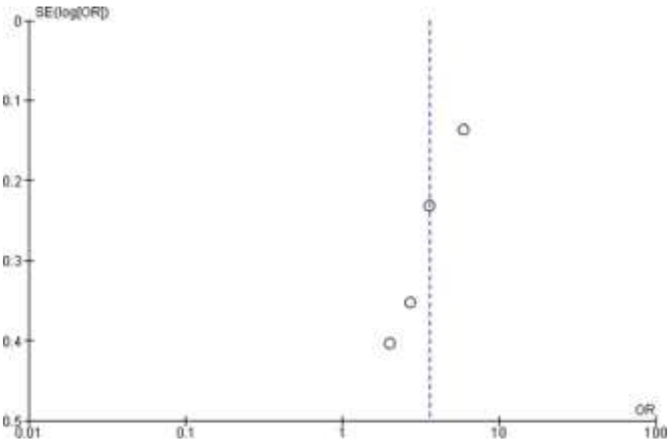

**Supplementary Figure S9**:**Publication Bias Assessment.** The funnel plot for mechanical ventilation analysis shows relatively symmetric distribution of the four included studies, with points clustered around the pooled effect estimate. Despite moderate heterogeneity ($I^2$ = 73%, p = 0.01), the studies appear fairly evenly distributed,

suggesting minimal publication bias. The consistent direction of effect across studies of varying precision (SE 0.137-0.403) strengthens confidence in the association between mechanical ventilation and stroke risk in COVID-19 patients.

**Supplementary Figure S10**:

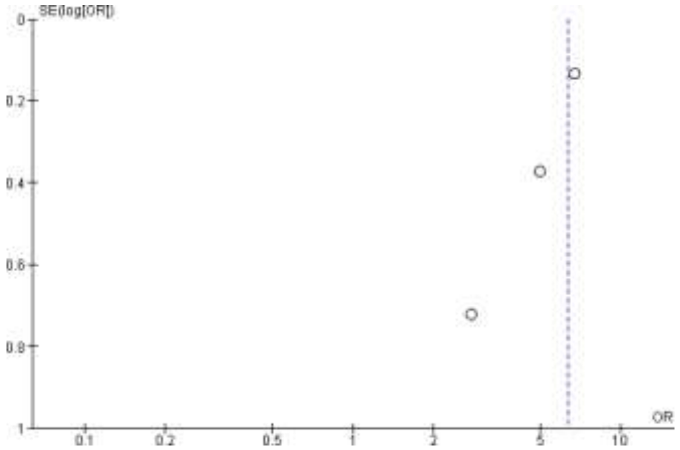

**Supplementary Figure S10**: **Publication Bias Assessment.** The funnel plot for ICU admission analysis shows relatively symmetrical distribution of the three included studies, with no evidence of substantial publication bias. The absence of heterogeneity ($I^2$ = 0%, p = 0.38) and consistent effect direction across studies of varying precision (SE 0.1356-0.7214) supports the reliability of the association between ICU admission and stroke risk. The weighted contribution is dominated by one large study (85.6%), which anchors the estimate near the pooled effect.

**Supplementary Figure S11:**

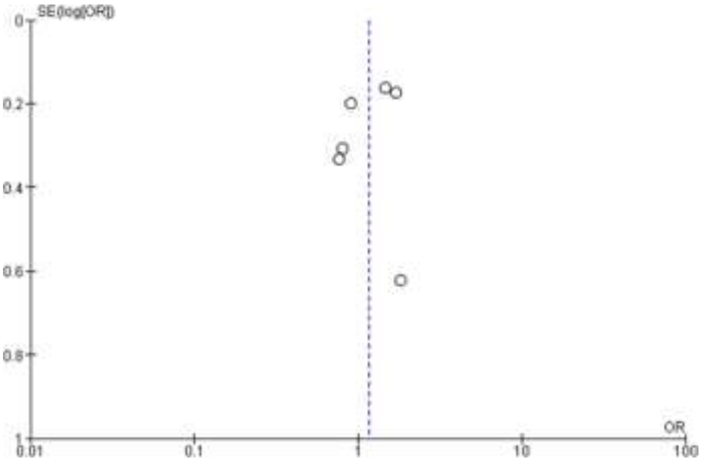

**Supplementary Figure S11: Publication Bias Assessment.** The funnel plot for gender analysis shows moderate asymmetry in the distribution of six studies, with points scattered on both sides of the null effect line. Despite moderate heterogeneity ($I^2$ =55%, p = 0.05), the studies appear relatively well-distributed across different precision levels (SE 0.164-0.622). The temporal pattern suggests evolving gender effects, with earlier studies (2020) showing higher odds ratios for males compared to more recent studies (2021-2023), though publication bias cannot be completely ruled out.

**Supplementary Figure S12:**

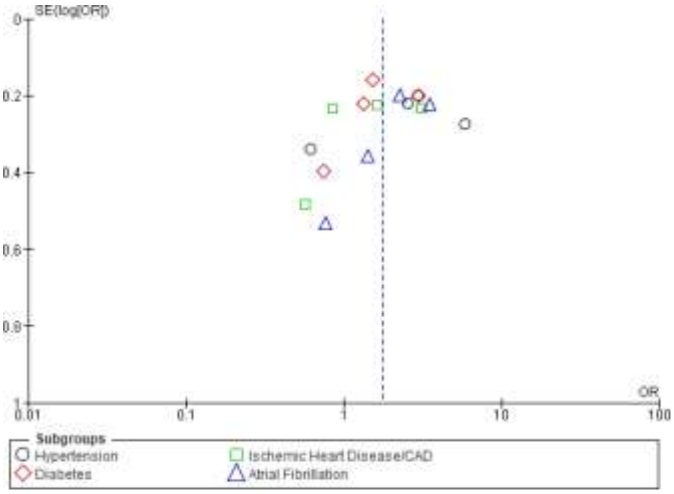

**Supplementary Figure S12**: **Publication Bias Assessment.** The funnel plot for comorbidity analysis shows the distribution of studies across four subgroups (hypertension, diabetes, ischemic heart disease/CAD, and atrial fibrillation), represented by different symbols. Despite high overall heterogeneity ($I^2 = 82\%$, $p < 0.00001$), the studies appear relatively evenly distributed across precision levels within each subgroup. The symmetrical pattern around the null effect line for each comorbidity suggests minimal publication bias, though heterogeneity varies by subgroup (hypertension: 89%, diabetes: 78%, IHD/CAD: 85%, atrial fibrillation: 68%). The consistent representation of studies across different precision levels strengthens confidence in the subgroup analyses.
